# Viral genetic variability in wastewater predicts changes in community infection levels

**DOI:** 10.1101/2025.10.24.25338735

**Authors:** Dustin T. Hill, Rafael Schulman, Ian Vasconcellos Caldas, Christopher Dunham, Yifan Zhu, Daryl Lamson, Lindsey Rickerman, Kirsten St. George, Yasir Ahmed-Braimah, Hyatt Green, Brittany L. Kmush, Frank Middleton, David A. Larsen

**Author notes:** Corresponding authors: Dustin T. Hill and David A. Larsen Department of Public Health Syracuse University Syracuse, NY 13244 USA (Email: DTH; DAL.

## Abstract

Sequencing viruses found in community wastewater facilitates the study of diversity in circulating viruses at the population level. By analyzing 12,290 wastewater samples collected between January 2023 and April 2025 in New York State, USA from 196 sampling sites across 57 counties, we assessed the diversity of the SARS-CoV-2 genome and how it changed over time compared to changes in COVID-19 infections and hospitalizations. We calculated three measures of SARS-CoV-2 genome diversity across all samples: nucleotide diversity (*π*), Shannon diversity (H), and viral variant count. We found that diversity increased with a rise in COVID-19 incidence and hospitalizations for all three measures (with a Spearman *ρ*> 0.8, p<0.001). The genetic diversity of the spike protein region had the highest correlation with the incidence of cases (*ρ* = 0.92, p<0.001 for *π, ρ* = 0.91, p <0.001 for H), and the statewide count of virus variants had a correlation coefficient of *ρ* = 0.85 (p<0.001) with case incidence. Additionally, the genetic diversity of the spike protein predicted 90.1 percent of the variance of COVID-19 case incidence. Our results demonstrate the potential for viral diversity analysis from wastewater in predicting epidemiological outcomes.

---

A COMMUNITY’S wastewater is a valuable source of public health information including circulating pathogens among the population. Routinely tested in the global polio eradication campaign (Asghar *et al*. 2014), wastewater testing also became a common method to track the spread of COVID-19 (Medema *et al*. 2020). The primary metric derived from wastewater samples to estimate infections has been the amount of pathogenic RNA or DNA recovered from a given unit or volume of wastewater sample, i.e. the viral load or concentration (Philo *et al*. 2021). Measuring amounts of pathogens in wastewater, independent of pathogen diversity, has been effective for estimating infection counts (Peccia *et al*. 2020), estimating the reproductive number (Hill *et al*. 2023), and determining areas free from disease (Larsen *et al*. 2022). However, determining viral concentration in wastewater is prone to problems stemming from differential shedding (Crank *et al*. 2022), sampling bias (Feng *et al*. 2021), persistence in sewage infrastructure (Li *et al*. 2023; Yang *et al*. 2022), and the inability to account for evolutionary changes of the virus that impact shedding profiles (Machkovech *et al*. 2024).

Diversity analysis of pathogen populations in wastewater has the potential to bypass some of these limitations. Wastewater populations reflect a mix of viral variants that are circulating in a community at the time of sampling (Fontenele *et al*. 2021; Izquierdo-Lara *et al*. 2021) and has been used to identify the spread of different lineages of SARS-CoV-2 (Bar-Or *et al*. 2021; Schenk *et al*. 2024). However, to date tracking SARS-CoV-2 variants through wastewater has been mostly descriptive, focused primarily on identifying taxonomically classified lineages. As a single wastewater sample represents the entire community (Yousif *et al*. 2023), we reasoned that diversity measured in viral genomes from wastewater would reflect epidemiologic changes in virus transmission (Mandal and Mandal 2023).

While often estimated in human samples (McCrone and Lauring 2016), genetic diversity measures the variation in DNA sequences in a population, and can be applied to a wastewater sample. The variation in the genetic material in a wastewater sample should reflect the variation of the virus in the community. As a virus spreads through a host population, viral genomes accumulate mutations during within-host evolution, some of which are propagated onwards upon transmission to subsequent hosts. During times of increased cases, the viral genome undergoes increased replication, thereby producing more mutations. We hypothesize that this increase in mutations will be reflected by an increase in diversity in wastewater-derived viral genomes and correlate with higher rates of infection. Moreover, mutations detected in wastewater will be observed in specific regions of the SARS-CoV-2 genome that reflect the ongoing biological process generating diversity in the viral population. For SARS-CoV-2 the regions of highest interest have been the spike protein due to its role in cellular transmission (Guruprasad 2021; Zhang *et al*. 2021) and the open reading frames (ORF) region due to its role in replication. For example, the ORF1a/b region encodes the 3C-like protease (NSP5), which is the target of treatments such as Paxlovid (nirmatrelvir or ritonavir) (Cho *et al*. 2023). In contrast, mutations in the spike protein of SARS-CoV-2 led to significant evolutionary shifts in the virus with the most prominent being the mutation of the virus from the Delta to the Omicron variant in 2021 (Kumar *et al*. 2022). Mutations in each of these regions present potential evolutionary benefit to the virus and we expect higher diversity in each of these regions of the SARS-CoV-2 genome in wastewater samples.

We employed three methods for estimating diversity in wastewater samples, drawing from population genetics, information science, and taxonomy-based approaches. 1) Diversity in a wastewater sample using per-base nucleotide diversity (*π*), defined as one minus the sum of squares of allelic frequencies across the viral genome (Tajima 1983). Using a modified version of *π*, we estimate the variation for each sequenced wastewater sample, hereafter referred to as *π*_ww_. 2) Shannon’s H (also called Shannon diversity, Shannon-Wiener index, or Shannon entropy) is another common measure of diversity and can be calculated per sequenced wastewater sample, hereafter H_ww_ (Keylock 2005). Both *π*_ww_ and H_ww_ are calculated per-base in each sequenced wastewater-derived viral genome, and a single per-sample score is calculated by aggregating per-base scores across the genome. In addition to these two aggregated per-base diversity measures, we also implemented a lineage-based diversity calculation. To do this, we use the deconvolution software Freyja to infer the number of frequencies of distinct named lineages that are identified in each sample, with more lineages implying higher diversity (Karthikeyan *et al*. 2022). Using whole genome sequences from 12,290 wastewater samples collected across New York State (NYS) spanning a 28-month period, we now show that nucleotide diversity of the SARS-CoV-2 genome as measured from wastewater predicts COVID-19 infections and hospitalizations.

## Results

From across New York State we sequenced a total of 12,290 wastewater samples between January 1, 2023 and April 20, 2025 (Table S1) that are included in the present study. These samples were collected from untreated wastewater at 196 unique sampling locations in 57 counties (Figure 1). Sequenced samples had all tested positive for SARS-CoV-2 with a mean PCR cycle threshold (Ct) value of 37. Sequenced samples had a mean depth of 450 reads and a mean coverage of 90 percent (range across all samples of 53 to 99 percent). The depth of the sequencing read was correlated with the Ct value of the PCR test, and depth had a weak log-linear relationship with SARS-CoV-2 RNA concentration S1 as other researchers have observed for viruses (Bergner *et al*. 2025).

**Figure 1:**
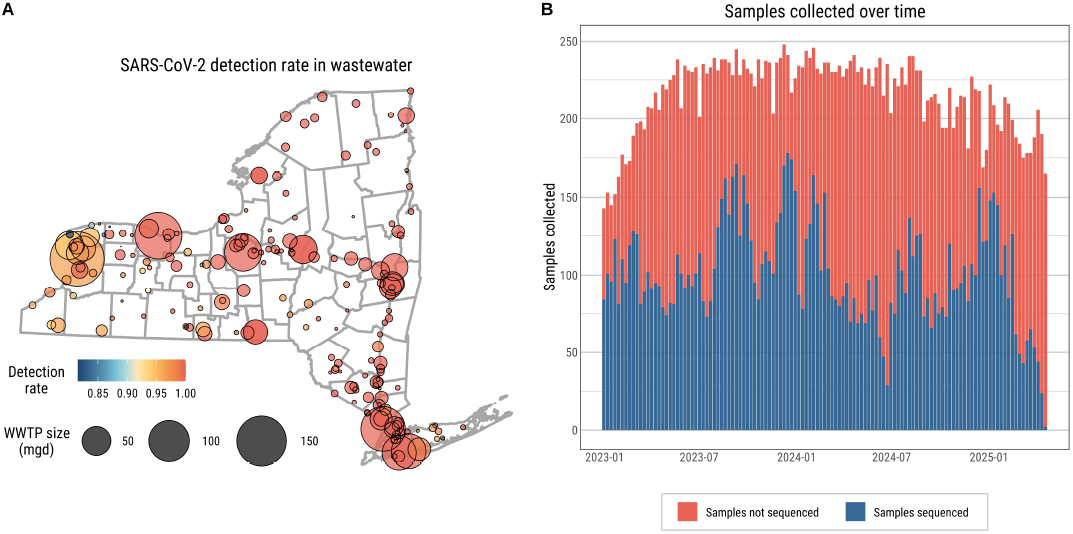
Study area with sample locations, rate of detection, and total samples collected over time. A) Wastewater sampling sites in NYS and total SARS-CoV-2 detection rate per site. B) Samples collected over time for SARS-CoV-2 quantification. All samples were sent for sequencing. Successful sequences were reported; sequences that were unsuccessful lacked enough genetic material to produce adequate results.. Data from 12,290 successful sequences were reported and analyzed herein.

We observed varying levels of genetic diversity across the SARS-CoV-2 genome over time. Specifically, the ORF 1a non-structural proteins 5 and 6 (NSPs 5 and 6) showed high diversity for both *π*_ww_ and H_ww_ (Figure 2). Genetic diversity was also high relative to the rest of the genome in ORF 1b for NSP 16, also known as 2’-O-methyltransferase (hereafter 2’ O-Mtase), and in the spike protein (Figure 2). Within the spike protein, two regions showed higher diversity than the rest of the spike: the S1 N-Terminal Domain (S1-NTD) and the S1 Receptor Binding Domain (S1-RBD). These regions showed consistently higher diversity over time than other parts of the genome but still varied temporally (Figure 2a and Figure 2b). These regions also showed high *π*_ww_ across wastewater samples (Figure 2c) and high Shannon H_ww_ (Figure 2d).

**Figure 2:**
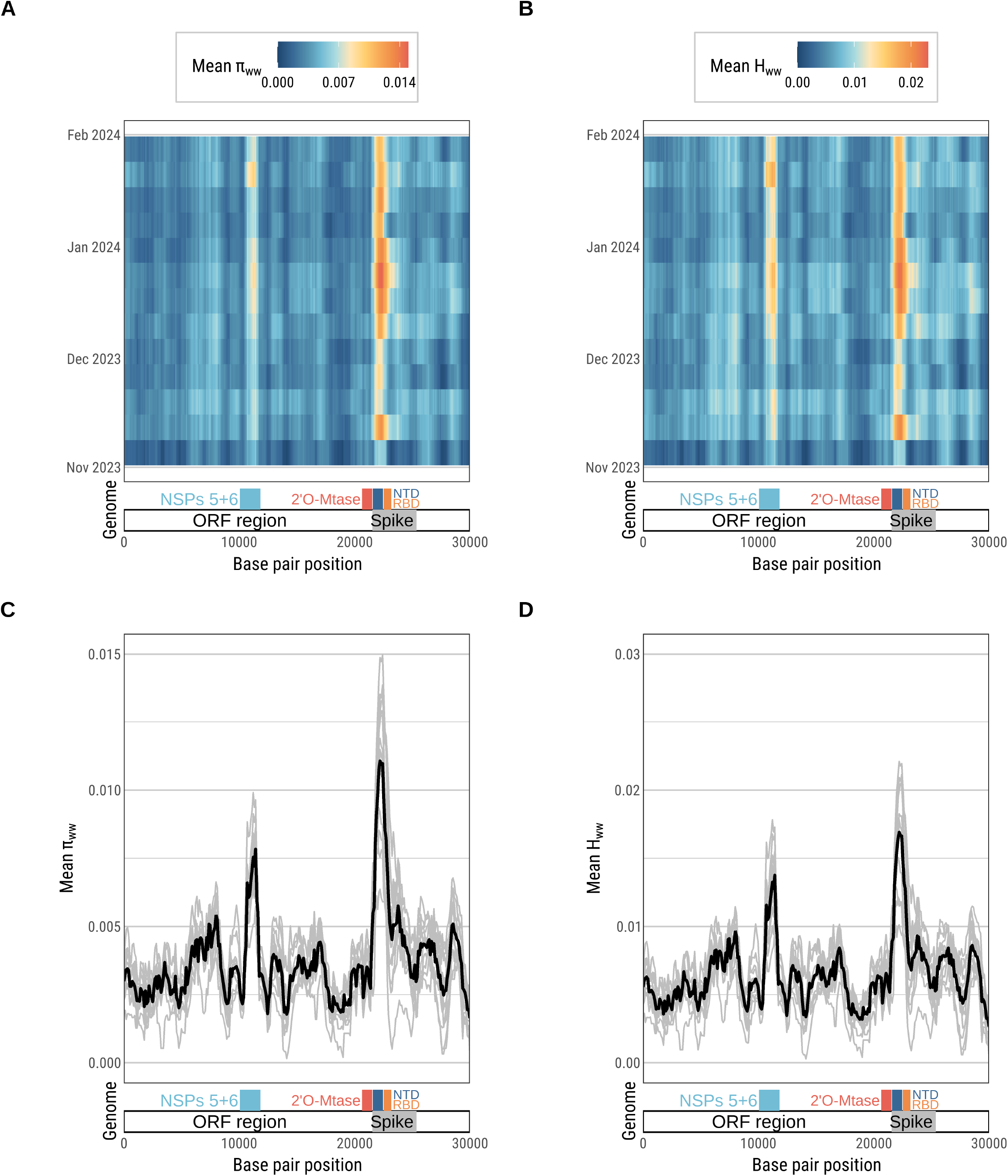
Nucleotide diversity of SARS-CoV-2 from a single wastewater treatment plant November 2023 – February2024. Genetic diversity varied across time and across the genome. The highest genetic diversity is observed for the NSP 5 and 6 region and the spike protein region in January 2024 as measured by both *π*_ww_ (A,C) and H_ww_ (B,D). In Figure 2C and 2D each sample is represented by a grey line with the thicker black line indicating the mean across the samples over this time period.

After identifying regions of high diversity in sequenced SARS-CoV-2 genomes, we examined these regions of the genome across samples throughout NYS over the entire study period, weighting each sample proportional to the population it represents. We also included the total number of reported virus variants from Freyja. The S1 NTD showed the strongest correlation with case incidence across time and across samples (Figure 3). This association was found for both genome diversity measures, *π*_ww_ and H_ww_ as well as virus variant counts, which increased as cases increased (Figure 3). Similar associations were also observed at county and regional spatial scales (Figure S2, Figure S3).

**Figure 3:**
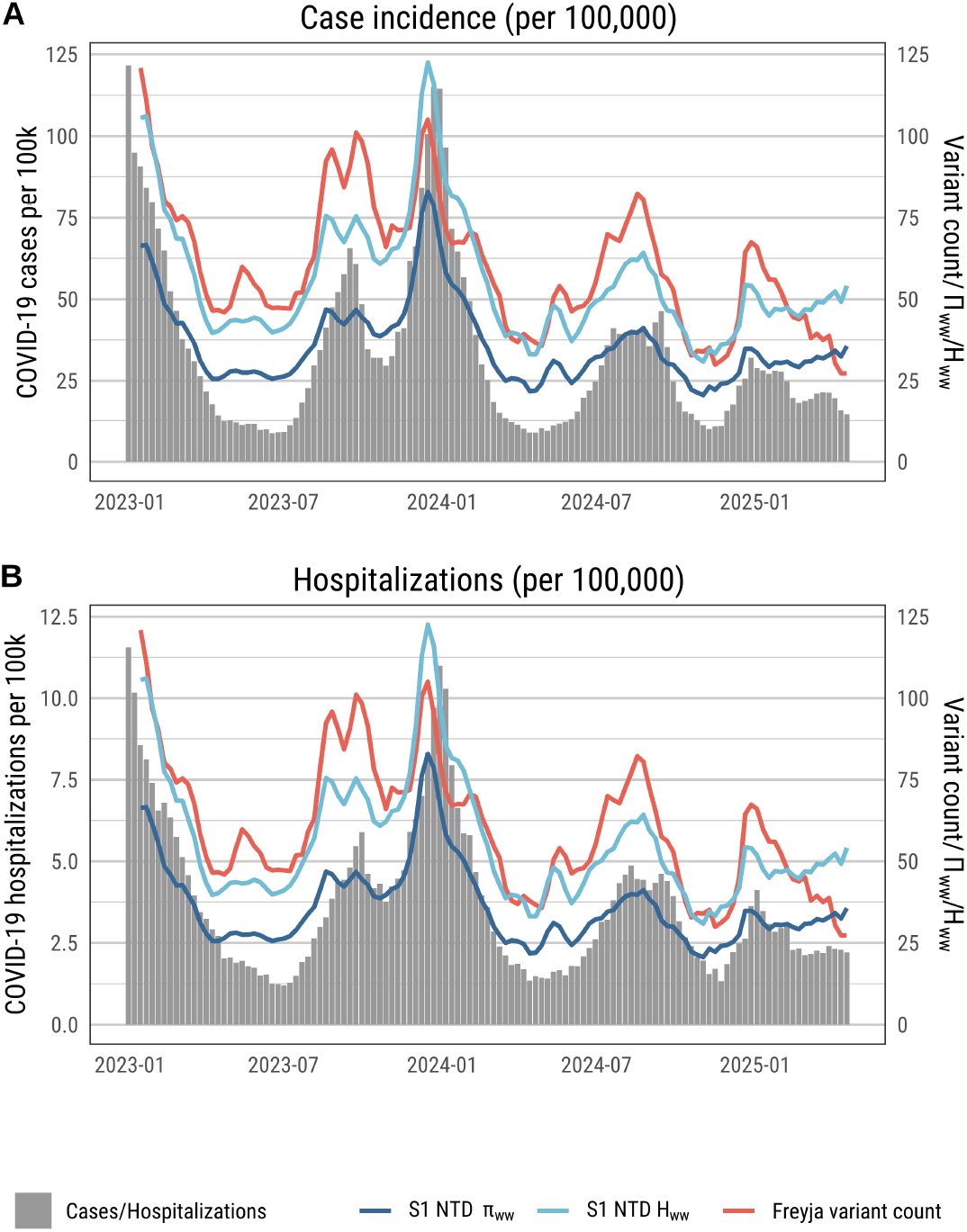
Diversity measures across all sewershed samples compared to clinical data. A) The left y-axis measures COVID-19 case incidence per 100,000. The right y-axis measures Freyja variant count, S1 NTD *π*_ww_ * 10,000, and S1 NTD H_ww_ * 10,000. All three diversity measures are displayed over time as population weighted 3-week rolling averages across all sewersheds. B) The left y-axis measures COVID-19 hospitalization incidence per 100,000. The right y-axis measures Freyja variant count, S1 NTD *π*_ww_ * 1,000, and S1 NTD H_ww_ * 1,000. All three diversity measures are displayed over time as population weighted 3-week rolling averages across all sewersheds.

We next compared the Spearman correlation between each region of the genome’s *π*_ww_ and H_ww_ values with COVID-19 case incidence and hospitalization incidence per 100,000 population, with sample weight proportional to the size of the population represented. While several regions of the genome showed high diversity and correlation with clinical data (e.g., NSP 5 and 6, S4, S5), it was the spike region that had the highest correlations (full correlations reported in S2). Specifically genetic diversity of the S1 NTD region correlated with COVID-19 case incidence whether using either *π*_ww_ or H_ww_ (*ρ* = 0.92, p < 0.001) and COVID-19 hospitalizations (*ρ* = 0.87, p < 0.0001) (S2). The count of virus variants also correlated with COVID-19 case incidence (*ρ* = 0.85, p < 0.001) and hospitalizations (*ρ* = 0.81, p < 0.0001). The correlation at the state level for all three diversity measures (*π*_s_, H_ww_, and virus variant count) was higher than correlation between statewide virus concentration in wastewater (Figure 4). Fisher’s Z-transformation test showed that S1 NTD *π*_s_ correlation was significantly different from the concentration for correlation (Z = 5.087, p <0.01).

**Figure 4:**
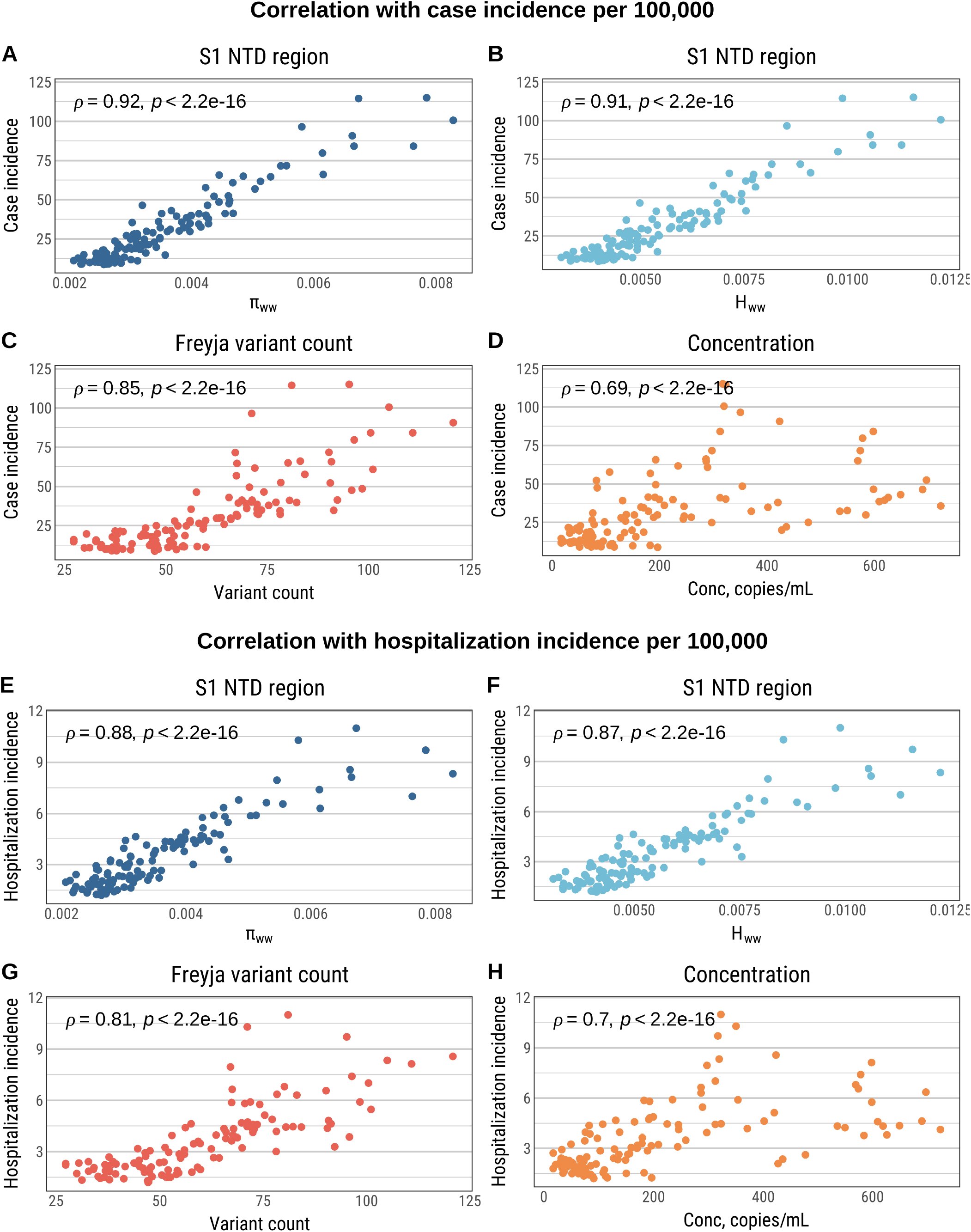
Relationship between clinical measures of COVID-19 and values derived from wastewater. A) S1 NTD *π*_ww_ and COVID-19 case incidence, B) S1 NTD H_ww_ and case COVID-19 incidence, C) Freyja variant counts from wastewater and COVID-19 case incidence, D) concentration of SARS-CoV-2 RNA in wastewater and COVID-19 case incidence, E) S1 NTD *π*_ww_ and COVID-19 hospitalization incidence, F) S1 NTD H_ww_ and COVID-19 hospitalization incidence, G) Freyja variant counts from wastewater and COVID-19 hospitalization incidence, H) concentration of SARS-CoV-2 RNA in wastewater and COVID-19 hospitalization incidence.

From the three measures of genetic diversity, we see strong correlation with the clinical data that are higher than the correlation based on the concentration of SARS-CoV-2 RNA in wastewater. We also see some possible early warning in the time series (Figure 3). Exploring the lead time of this relationship, we can see that the peak correlation between the measures of genetic diversity and COVID-19 incidence showed no lead time, but we did observe 1-week lead time for peak correlation of the diversity measures and hospitalizations across all weeks (Figure 5). All genome diversity measures showed early warning ranging between 1 and 2 weeks (S6), similar to the early warning proscribed to concentration data (Ahmed et al. 2021).

**Figure 5:**
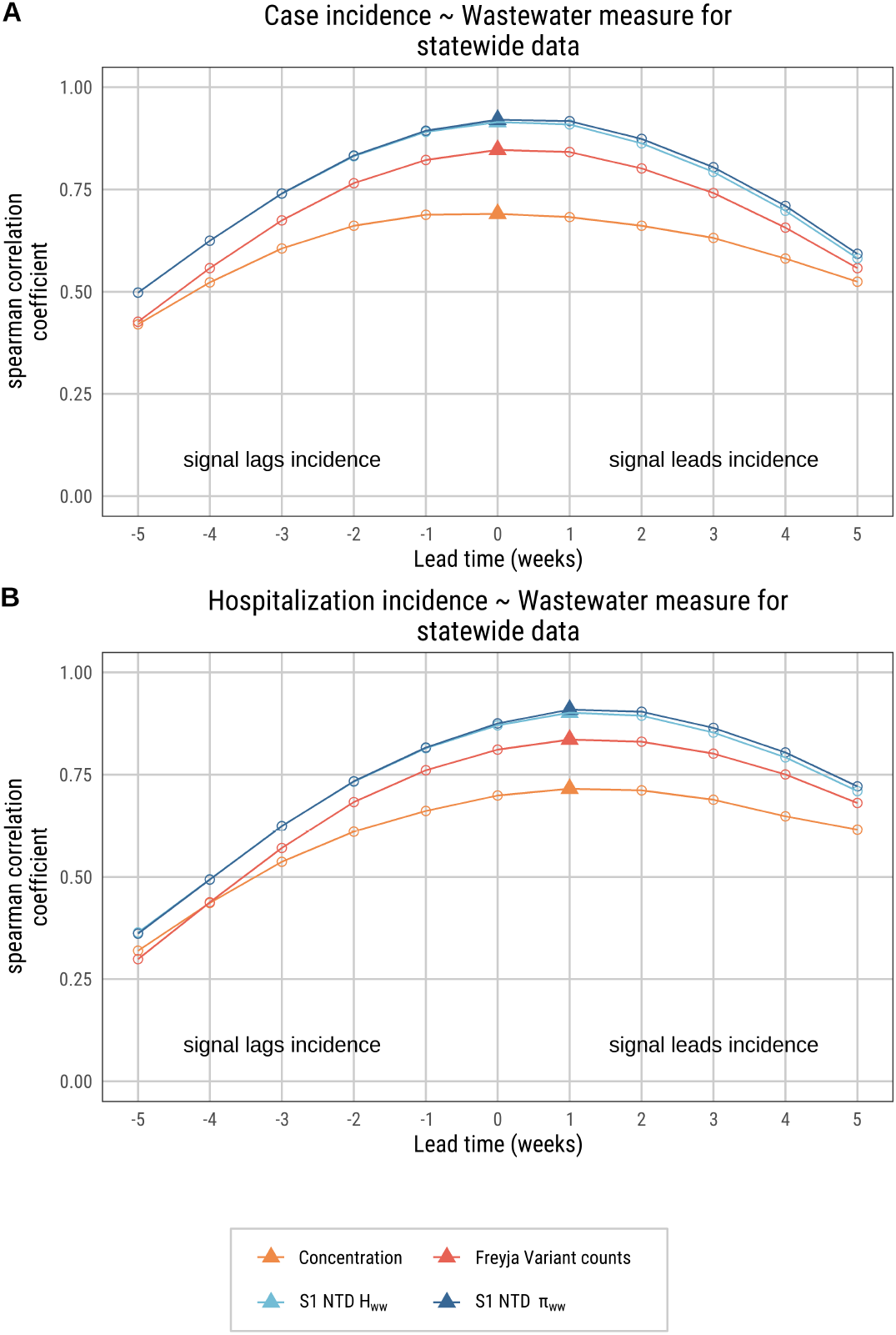
Change in Spearman correlation for diversity measures when the data are lagged to test for early warning potential. A) Spearman correlation for diversity measures/ concentration data and COVID-19 case incidence from statewide wastewater data. Peak lag is indicated by the triangles. The highest correlation for each measure was at time 0 suggesting little if any early warning for case incidence data with a weekly aggregation. B) Spearman correlation for diversity measures/ concentration data and COVID-19 hospitalization incidence from statewide wastewater data. Peak lag is indicated by the triangles. The highest correlation for each measure was when wastewater data were lagged 1 week suggesting early warning for hospitalization data with a weekly aggregation.

In a regression model, the explained variance of case incidence by S1 NTD *π*_ww_ was much higher than concentration with an R^2^ of 0.901 as compared to 0.376 for concentration (S3). In addition, the standardized multivariate effect size for S1 NTD *π*_ww_ was much higher than for concentration (*β*=20.206 v. *β*=3.709) even when including depth, number of samples collected, and coverage as covariates. These same patterns in generalized linear mixed models were observed for county and regional aggregations of the diversity data (Table S4, Table **??**). Additionally, the model shows that one standard deviation increase in the S1 NTD *π*_ww_ value was associated with an increase in incidence by 20.206 cases per 100,000 (Table S3).

Variance explained is an important indicator of the predictive ability of the diversity measures, but to determine if diversity can operate in a forecasting role, we also tested for Granger causality to see if one time series (diversity) could predict the second time series (clinical data). Each diversity measure was highly predictive of both case incidence and hospitalization incidence (Figure S6). This suggests that the diversity measures could be used to forecast unobserved hospitalization incidence because there is likely lead time in the lagged diversity measures. There was also no evidence of bidirectional causality suggesting that lagged clinical data would not predict diversity in wastewater samples (Table S6).

Examining Freyja variants output without applying a threshold for abundance is a non-traditional approach to this kind of sequencing data. Thus, we also ran the analysis with a threshold for variant abundance and grouped by lineage family as is recommended. This resulted in a weak, negative correlation (*ρ* = -0.19, P value = 0.038) between the Freyja lineage counts and clinical data (Figure S7) and shows that during the peak surge in COVID-19 cases, one variant became dominant in a “selective sweep”. Using the five percent threshold for virus variant counts in their disaggregated form yielded a moderate negative correlation (*ρ* = -0.42, P value < 0.001) with the clinical data (Figure S8).

## Discussion

Our results show that measures of viral diversity from wastewater samples can be strong indicators of changes in community transmission levels, superior to the measured amount of viral nucleic acid from wastewater based on PCR. Genome-wide diversity of SARS-CoV-2 changed in parallel with changing transmission with certain portions of the genome showing higher diversity than other regions. The ORF 1a region for NSPs 5 and 6, the ORF 1b region for 2’-O-MTase, and the spike protein region S1 NTD all exhibited high genetic diversity, however, the S1 NTD region correlated the strongest with clinical data. The results suggest that there were a greater number of mutations in these genome regions at times when transmission was increased, and more people were infected.

For mutations in the SARS-CoV-2 genome to be passed on and increase in abundance, the mutations must exhibit a beneficial effect such as helping a virus evade antiviral drugs (Hedskog *et al*. 2010). Mutations in the NSP 5 and 6 regions would potentially hold benefits for the virus because these two proteins are part of the replication region, and NSP 5 is the target of recommended antiviral treatments (Lee *et al*. 2022). Similarly, 2’-O-MTase is also involved with replication, and mutations here have been linked with SARS-CoV-2’s ability to avoid immune responses (Deng *et al*. 2024)(Vithani *et al*. 2021). While both regions exhibited high nucleotide diversity (Figure 2), their correlations with clinical data were not as strong as S1 NTD’s correlation (Table S2, Figure S4). The spike protein sequence encoded within the S1 NTD and adjacent RBD region are essential for coronaviruses to attach to a host cell and infect the host organism (Zhang *et al*. 2021). Mutations in this region could be beneficial to the virus if it increases the chance of infecting a new host or avoiding host neutralizing antibodies (Kumar *et al*. 2022). (Kumar et al. 2022). Further, the increase in diversity of spike region S1 NTD in the wastewater samples follows the clinical data closely in our study, suggesting that there are more mutations present in this region in wastewater sequences when there are more infections in the community (Nelson and Hughes 2015).

Thus, we find that diversity of a pathogen in wastewater is a good measure of infections. When a surge in the number of infections is occurring, this is sometimes led by one virus lineage, such as we see in the data during the 2023-2024 winter season (Figure S7). The viruses that are being shed carry enough similarity to be in the same lineage family, which “takes over” in a “selective sweep”, thereby decreasing the number of lineages (Harris *et al*. 2018). At the same time, nucleotide diversity increases because of the high infection rate, and high viral mutation rate among the “sweeping” strain; even very small genome differences that would be classified as the same variant will increase the nucleotide diversity. While a “selective sweep” is a reduction in the overall number of virus lineages circulating (Boyle *et al*. 2022), our findings suggest that when a “selective sweep” occurs there is an increase in transmission and subsequently an increase in the overall nucleotide diversity of the virus as measured by *π*_ww_ or H_ww_. There are more infections and more unique virus variants circulating but without enough mutational differences to be considered a different lineage, resulting in greater overall genetic diversity of the virus even when fewer lineages are reported. Notably, our use of Freyja is different than what is recommended in that we include variants with very small prevalence levels (less than one percent). While Freyja does not recommend this for grouping variants or estimating prevalence, we avoid drawing any conclusions about variant prevalence and only focus on the diversity of the variants identified.

Additionally, we find that diversity is an indicator of disease transmission and an improvement over other measures, like virus concentration in wastewater (Figure 4, Figure 5). While it might be tempting to use a different measure from sequence data like depth of read as the indicator of the number of infections found in the sample, depth did not show the same strength of correlation with clinical data (Figure S9). Depth might also be obscured at times by single infections producing multiple reads of the virus (Balmer and Tanner 2011). Thus, diversity is a better indicator of transmission than depth of read.

Lastly, although we use SARS-CoV-2 as a model pathogen, we hypothesize that our findings may be generalizable to other pathogens. Similar work has been done using clinical studies of malaria (Gwarinda *et al*. 2021) and influenza (Croze and Kim 2021), where higher diversity occurs during periods of higher transmission. These results provide a new way to conceptualize and work with next-generation sequence data from wastewater samples beyond lineage-based analyses and underscore the utility of calculating a diversity measure, like *π*_ww_ or Shannon, since they are simple and not computationally intensive. The Freyja variant count is also a simple measure with similar effectiveness and utility.

Our study has some limitations. All studies using next generation sequencing data face potential issues from machine error, alignment errors, sequencing errors, and in our case, noise from the wastewater samples and PCR errors. Our labs conduct quality control and validation, and along with our statistical tests and robustness checks, we sought to limit these errors as much as possible. Also, during the study period, reporting of COVID-19 infections likely changed as testing patterns can shift between seasons. We used countywide case numbers, not geolocated infections to the sewershed, which provides an imperfect match for linking infections to wastewater data. This misalignment may be inconsequential, as wastewater treatment plants serve as sentinel surveillance sites for their surrounding region (Reckling *et al*. 2024) (Yu *et al*. 2024). Also, not all samples could be successfully sequenced, and our findings only represent the results of samples with successful sequencing. It is possible that excluded samples contain useful data that is missing from our study. We do not expect that this exclusion had a large impact on the overall findings given the number of samples that were sequenced and the long timeframe of the study. In addition, there might be concern that we are simply measuring depth of read in each sequenced sample where more diversity is present because there are a greater number of virus strains in the sample. This is unlikely because depth of read, while correlated with diversity, did not explain the variation in diversity (14 percent of the variation in *π*_ww_ was explained by depth, S10, S7), and random subsampling of reads of similar depth yields the same overall findings (S11). Further, diversity was more predictive of case incidence than depth in our GLMM. Diversity measures proved better indicators of transmission than concentration or depth suggesting that total virus levels in wastewater does not fully explain the associations that we are seeing.

Throughout the COVID-19 pandemic, wastewater genetic sequence data has been essential for tracking mutations of virus and detecting named variants of concern (Bar-Or *et al*. 2021). Building on these foundations, quantifying the diversity of viruses in a wastewater sample has the potential to improve predictive measures of new infections in a community. This offers a new direction for wastewater-based epidemiology, one that needs to be explored further.

## Materials and Methods

### Setting

New York State has been testing for SARS-CoV-2 in wastewater samples since May of 2020 (Neyra *et al*. 2023). Whole genome sequencing of SARS-CoV-2 was implemented statewide in 2022. A total of 196 sites were included in this study that had at least one valid whole genome sequence for a wastewater sample (Figure 1). New York City is considered a separate CDC jurisdiction, and they are not included in these analyses.

### Wastewater sample collection and processing

Wastewater samples were processed and analyzed for SARS-CoV-2 by five laboratories each with different methods (see S8 for detailed descriptions). Briefly, Quadrant Laboratories (Syracuse, NY) processed 9,276 samples using ultracentrifugation with a sucrose cushion and quantified SARS-CoV-2 concentration using reverse transcription quantitative polymerase chain reaction (RT-qPCR). Full documentation of these methods were previously published (Wilder *et al*. 2021). Wadsworth Center began processing samples in January 2025 using CERES Nanotrap and digital droplet PCR and processed 236 samples. University at Buffalo (SUNY Buffalo) processed 1,335 samples mostly for Erie and Niagara Counties and their processing Nanotrap® Enhancement Reagent 1 (Ceres Nanosciences) and Nanotrap® Microbiome A Particles (Ceres Nanosciences). SUNY Buffalo quantified SARS-CoV-2 N gene (Lu *et al*. 2020) using RT-qPCR. The fourth laboratory was located at SUNY Stony Brook, which processed 1,323 samples for this study from Long Island (Nassau and Suffolk counties). SUNY Stony Brook used polyethylene glycol (PEG) precipitation to process samples and quantified using digital PCR. The fifth and final laboratory was run by Genesee and Orleans County Health Department (GO Health) and they processed 196 samples for Genesee and Orleans Counties. GO Health used Innovaprep with ultrafiltration for processing and GT digital PCR for quantification. Further details for each lab and method are provided in the supplementary material. Descriptive statistics for concentration are reported in S9.

### Sequencing of wastewater samples

Whole-genome sequencing for SARS-CoV-2 was piloted in the fall of 2022 with sequencing of all samples beginning in December 2022. All five regional sequencing laboratories used the same methods. Upon receipt of samples, 25 µL are loaded onto the Genexus Integrated Sequencer as per the manufacturer’s guidelines (Thermo Fisher Scientific, Waltham, MA, USA). Whole genome sequencing is performed using the Ion AmpliSeq™ SARS-CoV-2 Insight Research Assay GX (Catalog #: A51307). The assay kit requires two assay files and the analysis plugins to be downloaded from Thermo Fisher™ Connect. Depending on the concentration of the samples, either the SARS CoV 2 Insight LowTiter Research Assay (< 1,000 copies) or SARS CoV 2 Insight Research Assay (< 1,000 copies) is implemented. Each assay performs library preparation, sequencing, and analysis. The raw sequence reads are mapped against the reference sequence (NC 045512.2) and trimmed, generating processed binary alignment map (BAM) files.

### Bioinformatic pipeline

Data from genetic sequencing (BAM files) were uploaded to Syracuse University from December 2022 to June 2024 and then Wadsworth Center from July 2024 to April 2025 for bioinformatic processing. The same processing pipeline was used by both institutions. Sequences were processed using a snakemake pipeline adapted from the Center for Food Safety and Applied Nutrition Wastewater Analysis Pipeline ((noa 2025)) to align and call SARS-CoV-2 variants. Sequence data were first filtered so they contained only reads that align to the SARS-CoV-2 Wuhan genome (accession number NC_045512.2). Alignments were run through QualiMap v2.2 (García-Alcalde *et al*. 2012)for quality control, and any sequences with less than 20x coverage across at least across 50% of the genome were discarded. The remaining sequences were run through Freyja v1.4 (andersen-lab/Freyja 2025) to identify COVID-19 lineages. The sequences went through variant calling and demixing, such that the output of this process was the relative abundance of each detected lineage in each sequence. Lineages are obtained from the reference set of UShER tree barcodes (Turakhia *et al*. 2021). Before every run, the set of barcodes used in the pipeline was updated to the latest available version, so that the results reflected the latest consensus on variant composition of lineages.

### Clinical data

COVID-19 hospital admissions and hospital incidence data were obtained from the NYS DOH COVID database (**?**). Data were reported during weekdays and were summed from the reported hospitals to the corresponding counties per week to get a total hospital admissions value and incidence per 100,000 population per week per county. Regional and statewide totals were similarly summed. COVID-19 case data and case incidence were also obtained from the NYS DOH COVID database (**?**) and these data were summed to the weekly level per county. Descriptive statistics are reported in S10.

### Viral diversity methods

Nucleotide diversity (Pi or) was calculated on each Freyja variants file for SARS-CoV-2 wastewater reads. First, iVar was run on BAM files to call variants, then they were filtered to samples that passed quality control. For each position of the reference genome with called variants, we calculated, which is the probability that two randomly chosen reads spanning that position are different. The equation is written as:

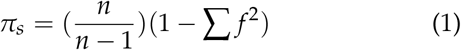

Where is the total number of reads spanning that position, is the frequency of a variant, and the sum over all variants at that position, thus providing a *π* per base, or *π*_b_. *π* is an established measure of nucleotide diversity. Tajima (Tajima 1983) published a review and discussion of how the measure relates to evolution of DNA sequences in populations.

Once obtaining the single value for per base, we continued into a second step to calculate the average diversity over genomic windows of a fixed size,, to reduce noise using the following equation:

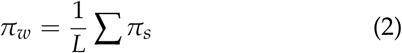

Where is the window size of base pairs, positions with no variation in the sample are considered to have equal to zero. Windows of 1000 base pairs (bps) in length were used with every window starting every 100 bps. Window sizes of 500 bps and 2000 bps were also explored to ensure that the 1000 bp window size did not bias the results. The other window sizes yielded the same findings for high diversity in the genome regions of interest (S12). A final genome-wide was calculated by taking the average across the for each wastewater sample (hereafter referred to as *π*_ww_). Averaging across windows is common in other studies because it avoids overestimating when portions of the genome are missing(Konopiński 2023).

We produced a Shannon H value (Sherwin *et al*. 2017) for each base pair following the same approach we used for *π*_s_ using the following equation:

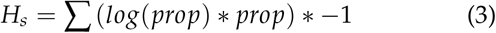

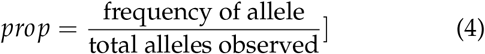

In addition to a diversity value for every sample for the entire genome, we also estimated diversity for specific regions of the SARS-CoV-2 genome (S11). These regions were selected for individual analysis because they each showed high diversity relative to the rest of the genome across wastewater samples (Figure 22) and have biological significance to SARS-CoV-2. For depth of read, we used the depth of the Freyja variant reads instead of the total sample depth. Freyja variant read depth was perfectly correlated with total sample depth suggesting the analysis would be the same if we were to use total sample depth (S13).

Last, using the BAM files and Freyja summary output, we counted the number of named variants (i.e., strains or lineages) found in each wastewater sample. We counted variants per sample from the total Freyja output and a second count using a threshold of 5 percent, where variants reported with abundance values below 5 percent were excluded. We repeated both measures for lineage families (groups of variants according to phylogeny).

### Statistical analysis

Genome diversity values were calculated for each sewer-shed that had a sample sent for sequencing each week and that passed quality control. If a sewershed had more than one sample in a given week, diversity values were averaged for that site for that week. The data were further aggregated to the county, region, and statewide level using the population of the sewershed as the weight. Most of the results in the main text use the statewide population weighted data with county and regional results reported in the supplement.

To compare the values to other time-series data, like COVID-19 cases and hospital admissions, a three-week right-adjusted rolling average was calculated for all measures (diversity, cases, etc.). This step smoothed some of the noise between sampling weeks and reporting weeks for clinical data. To test correlation between diversity and clinical data, Spearman’s rank correlation coefficient was used because many associations were non-linear.

Since the signal from wastewater concentration data are known to lead clinical cases and hospital admissions (Hill *et al*. 2023), we tested for a similar lead time associated with diversity. We used the Spearman correlation and tested for a change in correlation when diversity was moved backward in time, or forward in time, and we tested values from 1 to 10 weeks prior and post case reporting.

To estimate the variance explained by the diversity measures, we fit a series of generalized linear mixed models (GLMMs) for county, region, and statewide diversity and estimated the ability to predict COVID-19 cases and hospitalizations per 100,000 population. The GLMMs account for repeated measures using an autocorrelation correction for time. Further, we compared the variance explained by diversity to the variance explained by SARS-CoV-2 virus concentration in wastewater. Last, to determine the predictive ability of the diversity measures, we used Granger causality (Granger 1969), (Thurman and Fisher 1988). Granger causality is a measure of prediction, not causality, and determines if one time series can predict another time series by testing different lags. If Granger causality exists, then past values (lags) of variable X_1_ will predict future values of X_2_ with greater ability than lagged values of X_2_ alone. All statistical analyses were completed in R version 4.1.1. For significance tests, alpha was set to 0.05.

## Data availability statement

Wastewater concentration data are available to download from the NYS Department of Health at this link: https://health.data.ny.gov/Health/New-York-State-Statewide-COVID-19-Wastewater-Surve/hdxs-icuh/about_data. COVID-19 case data are available at this link: https://health.data.ny.gov/Health/New-York-State-Statewide-COVID-19-Testing/jvfi-ffup/about_data.

COVID-19 hospital admissions data are available at this link https://health.data.ny.gov/Health/New-York-State-Statewide-COVID-19-Hospitalizations/jw46-jpb7/about_data.

Sequencing results are available on NCBI for the BioProject for the NYS WWSN at this link: https://www.ncbi.nlm.nih.gov/bioproject/?term=PRJNA896

Bioinformatics pipeline. The wastewater sample processing pipeline can be accessed at this GitHub page: https://github.com/YazBraimah/NYWWS/tree/split

All code used to produce the analyses here are collected on a project GitHub page here: https://github.com/nys-wwsn/nucleotide-diversity. The GitHub includes all the data used to produce these analyses.

## Credit authorship

**DTH:** conceptualization, methodology, software, validation, formal analysis, investigation, data curation, writing – original draft, writing – review and editing, visualization. **RS:** conceptualization, methodology, software, validation. **CD**: data curation, conceptualization, writing – review and editing. **IVC:** conceptualization, methodology, software, validation, formal analysis, investigation, data curation. **YAB:** conceptualization, software, project administration, supervision, writing – review and editing. **LR:** data curation, software. **KSG:** Supervision, funding acquisition, writing – review and editing. **DL:** Data curation. **FM:** Supervision, writing – review and editing. **BK**: Conceptualization, writing – review and editing. **HG:** Supervision, conceptualization, writing – review and editing. **YZ:** conceptualization, writing – review and editing. **DAL:** conceptualization, methodology, resources, writing – review and editing, supervision, project administration, funding acquisition

## Acknowledgements

We would like to thank the New York State Wastewater Surveillance Network and the New Yok State Sequencing Consortium Labs for shipping and analyzing PCR and sequencing data. We also want to thank the wastewater treatment plant operators for collecting samples for analysis.

## Supporting Information

### Supplementary Methods

Wastewater samples were processed and analyzed for SARS-CoV-2 by five laboratories each with different methods (see Table S4 for brief descriptions). Briefly, Quadrant Laboratories processed 9,276 samples using ultracentrifugation with a sucrose cushion and quantified SARS-CoV-2 concentration using reverse transcription quantitative polymerase chain reaction (RT-qPCR). Full documentation of these methods were previously published (1). Wadsworth Center began processing samples in January 2025 using Nanotrap Magnetic Beads and Digital PCR. University at Buffalo (SUNY Buffalo) processed 1,335 samples mostly for Erie and Niagara Counties and their processing method took the 24h influent samples of 9.75 mL and mixed them with 100 µL of Nanotrap^©^ Enhancement Reagent 1 (Ceres Nanosciences) and 150 µL of Nanotrap^©^.Microbiome A Particles (Ceres Nanosciences). Viruses were separated from the wastewater using KingFisher Apex Benchtop Sample Prep system from Thermo Fisher. After separation, the nucleic acids were extracted using MagMAX Viral/Pathogen Nucleic Acid Isolation Kits (Thermo Fisher) then eluted in MagMAX Viral/Pathogen Elution Buffer (Thermo Fisher) and stored at -80°C. SUNY Buffalo quantified SARS-CoV-2 N gene(2) using RT-qPCR. The RT-qPCR quantification used 10 µL RT-qPCR reaction mixtures consisting of 5 µL of 2x iTag Universal Probes Reaction Mix from Bio-Rad, 0.25 µL of 50x iScript reverse transcriptase also from Bio-Rad, 0.75 µL of 2019nCoV N2 (RUO Kit, IDT), and 4µL of undiluted nucleic acid extracts. RT-qPCR of the nucleic acid extracts. The SARS-CoV-2 reactions were heated at 50°C for 15 minutes, 95°C for 1 minute, and 40 cycle of 95°C for 10 seconds and 60°C for 30 seconds. Each RT-qPCR assay was conducted in duplicates or triplicates on a CFX96 Touch Real-Time PCR Detection System (Bio-Rad).

The fourth laboratory was located at the SUNY Stony Brook, which processed 1,323 samples for this study from Long Island (Nassau and Suffolk counties). 24h composite samples of raw sewage were centrifuged at 4200 rpm for 30 min at 4°C to remove large particles and debris before polyethylene glycol (PEG) precipitation. Recovery rates were evaluated using bovine coronavirus (BCoV), which belongs to the same genus as SARS-CoV-2, was spiked into the supernatant. The viral particles in 40 mL of samples were precipitated with PEG 8000 (Millipore Sigma, Burlington, MA) and NaCl (5 M, Millipore Sigma, Burlington, MA) and then incubated overnight at 4°C. RNA from the PEG-precipitated wastewater was extracted by Qiagen QIAamp DSP viral RNA mini kit (Qiagen, Hilden, Germany) according to manufacturer’s instructions and eluted in 100 µ L by nuclease-free water. The concentrations of RNA were measured by NanoDrop One Spectrophotometer (Thermo Fisher Scientific, Waltham, MA). All RNA samples were stored at -80 °C and subjected to cDNA synthesis within the same day of RNA extraction to avoid losses associated with storing and freezing and thawing RNA extracts..

Quantification for SUNY Stony Brook samples was done using reverse transcription by High Capacity RNA-to-cDNA Kit (Applied Biosystems, Waltham, MA) at 37 °C C for 60 min, and stored at -20 °C until further analysis. The cycling condition was 95 °C for 5 s and 55 °C fr 40 s, and 98 °C for 10 min. The total volume of each reaction was 14.5 µ L containing 7.25 µ L of QuantStudio 3D Digital PCR Master mix v2 (Applied Biosystems, Massachusetts, USA), 0.725 µ L of TaqMan^©^ Copy Number Reference Assay RNase P (as an internal control, Applied Biosystems, Waltham, MA), 4.8 µ L of nuclease-free water, and 1 µ L of cDNA template. Digital PCR was performed using N1 primers and probe set from 2019-nCoV CDC EUA Kit (IDT # 10006606) and BCoV set against the BCoV gene as an external reference on a QuantStudio 3D Digital PCR (Applied Biosystems, Massachusetts, USA). Nuclease-free water was used as non-template control (NTC) and plasmids containing the complete nucleocapsid gene from 2019-nCoV (IDT # 10006625) were used as a positive control. Data analysis was performed with the online version of the QuantStudio 3D AnalysisSuite Cloud Software.

The fifth and final laboratory was run by Genesse and Orleans County Health Department (GO Health) and they processed 196 samples for Genesse and Orleans Counties. GO Health used Innovaprep Concentrating Pipette Select for ultrafiltration. A 1mL 10Tween 20 stock solution was added to the influent wastewater sample for every 100 mL of sample. Prepared sample was then pre-filtered using a 0.22 µ m prefilter. 125 µ L BCoV was added to each sample. Wastewater samples are maintained at 4 °C until processing. After filtration was complete, an Innovaprep Ultrafiltration PS 0.05 µ m Hollow Fiber Concentrating Pipette Tip (CPT) was connected to the Innovaprep CP Select. The Innovaprep elutes a wet foam into a 15 mL conical tube. The 15 mL tube was stored on ice until viral RNA extraction. Qiagen AllPrep Power Viral DNA/RNA Kit was used according to manufacturer’s instructions to extract viral RNA.

GO Health used the GT-Digital Influenza and SARS-CoV-2 Surveillance Multiplexed Assay kit (Fort Collins, Colorado) for Qiagen QIAcuity Digital PCR System. Manufacturer’s instructions were followed using this assay kit. In combination with the GT-Molecular kit for Influenza and SARS-CoV-2, GO Health used the Qiagen OneStep Advanced Probe Kit to create a Master Mix.The Master Mix was then placed into QIAgility a robotic workstation for an automated PCR setup; the machine automatically pipettes the appropriate amount of Master Mix in each sample on a 24 well nanoplate.The nanoplate was then run on the QIAcuity, where the concentration of each pathogen is determined.

**Table S1:** Table S1:Table: Descriptive statistics for genome-wide Pi.

| PCR Lab | Number of sampling sites | Number of samples | First collection week | Last collection week | Mean Pi | Min Pi | Median Pi | Max Pi | SD Pi |
| --- | --- | --- | --- | --- | --- | --- | --- | --- | --- |
| GO Health | 7 | 196 | 2023-02-05 | 2025-03-16 | 0.0028 | 0.00088 | 0.0028 | 0.0061 | 0.00110 |
| Quadrant | 149 | 9,200 | 2023-01-01 | 2025-04-20 | 0.0028 | 0.00048 | 0.0026 | 0.0190 | 0.00140 |
| SUNY Buffalo | 22 | 1,335 | 2023-01-22 | 2025-04-06 | 0.0034 | 0.00068 | 0.0032 | 0.0120 | 0.00140 |
| SUNY Stony Brook | 18 | 1,323 | 2023-01-01 | 2025-03-23 | 0.0032 | 0.00058 | 0.0028 | 0.0110 | 0.00170 |
| Wadsworth Center | 45 | 236 | 2024-12-22 | 2025-04-20 | 0.0028 | 0.00079 | 0.0029 | 0.0068 | 0.00095 |

**Table S2:** The correlation (Spearman’s R) between measures of genetic diversity and clinical COVID-19 measures.

| Region of the genome | COVID-19 case incidence |  | COVID-19 hospitalizations |  |
| --- | --- | --- | --- | --- |
|  | Pi<sub>ww</sub> | H<sub>ww</sub> | Pi<sub>ww</sub> | H<sub>ww</sub> |
| Genome-wide | 0.74 [0.63, 0.81] | 0.76 [0.66, 0.82] | 0.68 [0.56, 0.75] | 0.71 [0.60, 0.78] |
| NSPs 5 and 6 | 0.68 [0.58, 0.75] | 0.72 [0.63, 0.78] | 0.63 [0.51, 0.70] | 0.67 [0.57, 0.74] |
| 2' O-Mtase | 0.63 [0.50, 0.72] | 0.72 [0.62, 0.79] | 0.59 [0.45, 0.68] | 0.68 [0.56, 0.76] |
| Spike Protein | 0.82 [0.73, 0.87] | 0.82 [0.74, 0.87] | 0.75 [0.65, 0.82] | 0.77 [0.67, 0.83] |
| S1 NTD | 0.92 [0.88, 0.95] | 0.92 [0.87, 0.94] | 0.88 [0.82, 0.91] | 0.87 [0.81, 0.91] |
| S1 RBD | 0.63 [0.48, 0.74] | 0.67 [0.53, 0.76] | 0.61 [0.46, 0.70] | 0.64 [0.50, 0.72] |

**Figure S1:**
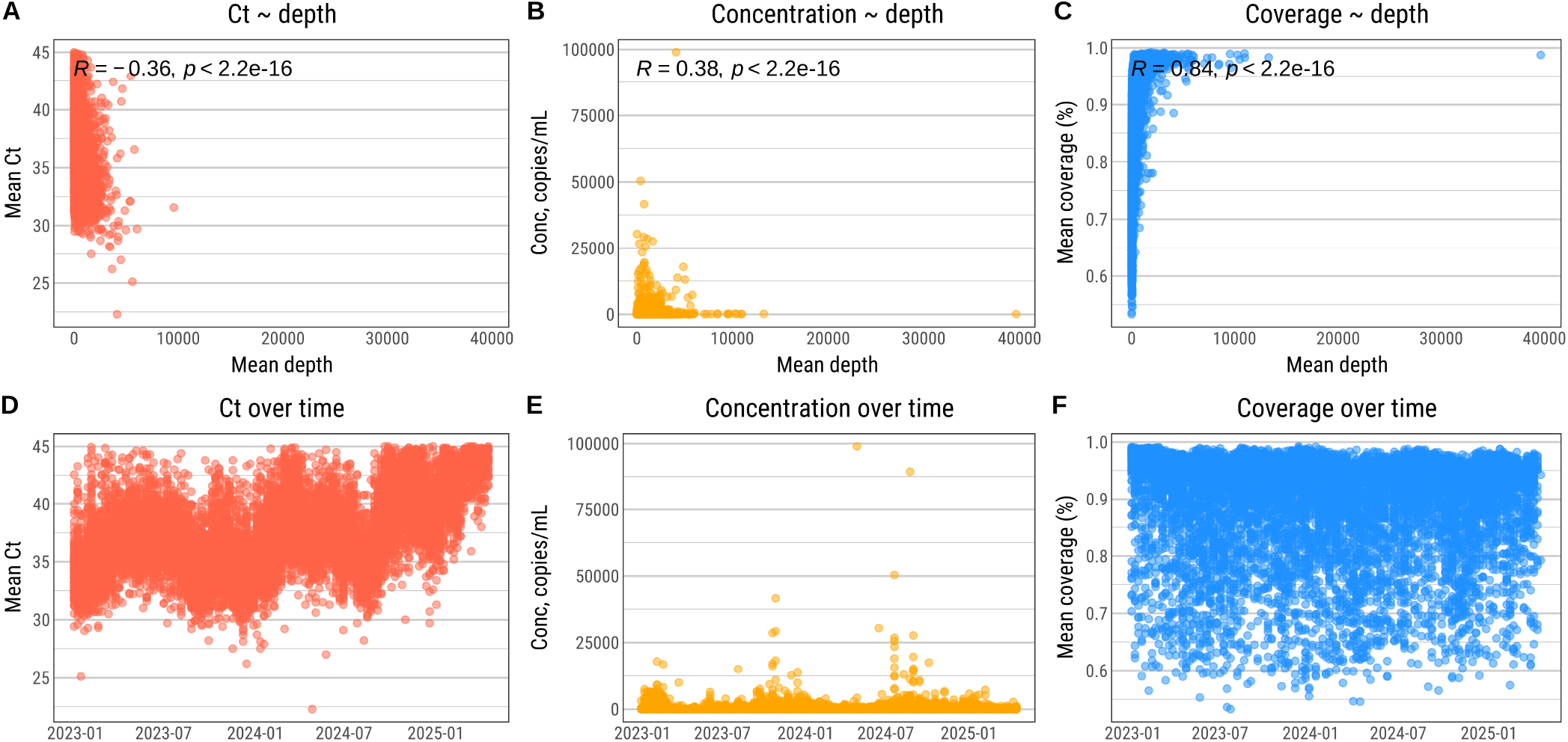
A) Scatterplot and Spearman correlation for coverage (Ct) and depth (number of reads). There is weak negative correlation and the association is non-linear. B) Scatterplot and Spearman correlation for concentration and depth. There is weak positive correlation and the association is non-linear. C) Scatterplot and Spearman correlation for sample genome coverage and depth. There is a positive, non-linear correlation for coverage and depth. D) Ct per sample over time. E) Concentration per sample over time. F) Genome coverage per sample over time.

**Figure S2:**
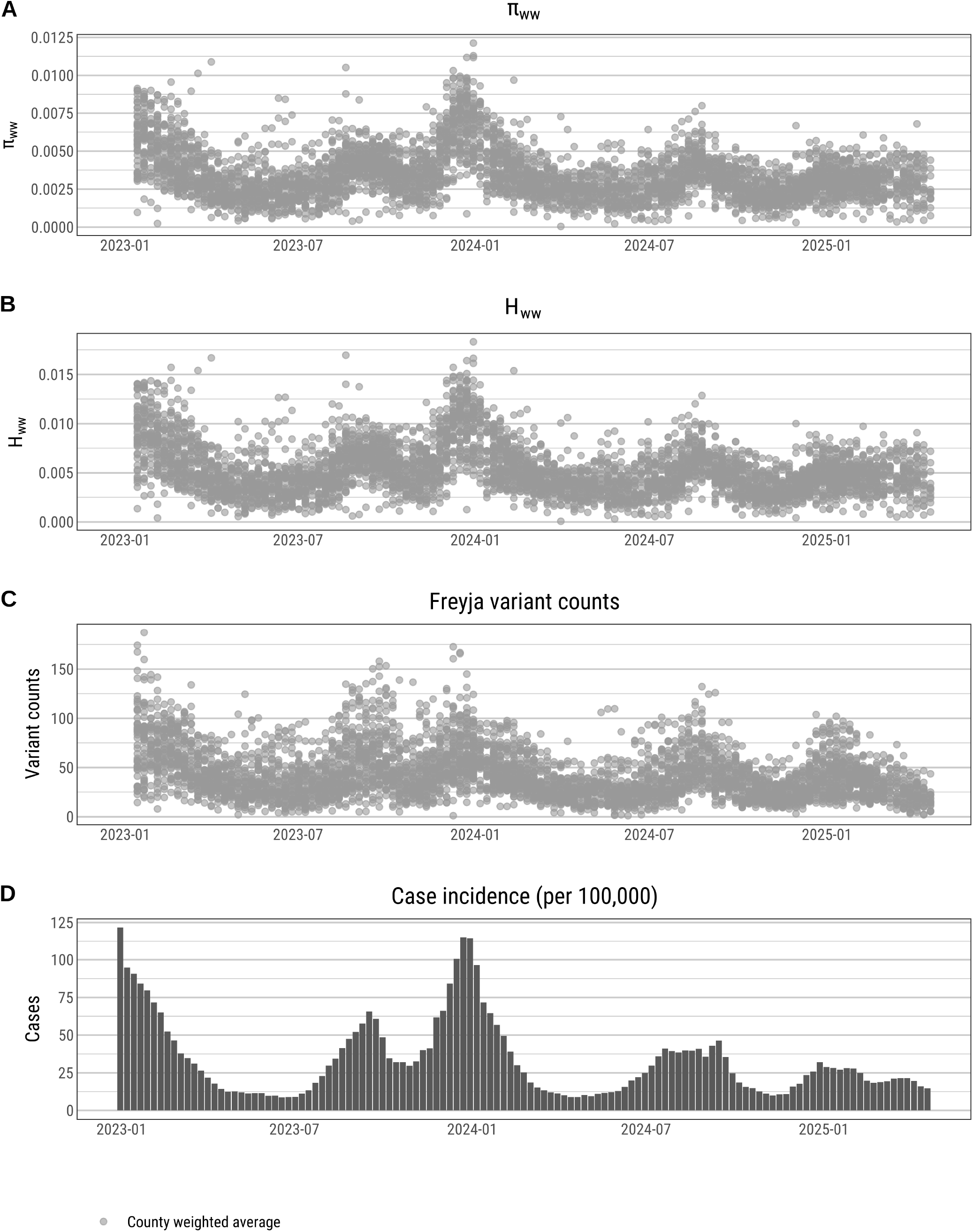
A) County population weighted average *π*_s_ estimates for the S1 NTD region per county, per week. B) County population weighted average H_s_ estimates for the S1 NTD region per county, per week. C) County population weighted average number of Freyja variant counts per county per week. D) Weekly statewide total cases per 100,000 population.

**Figure S3:**
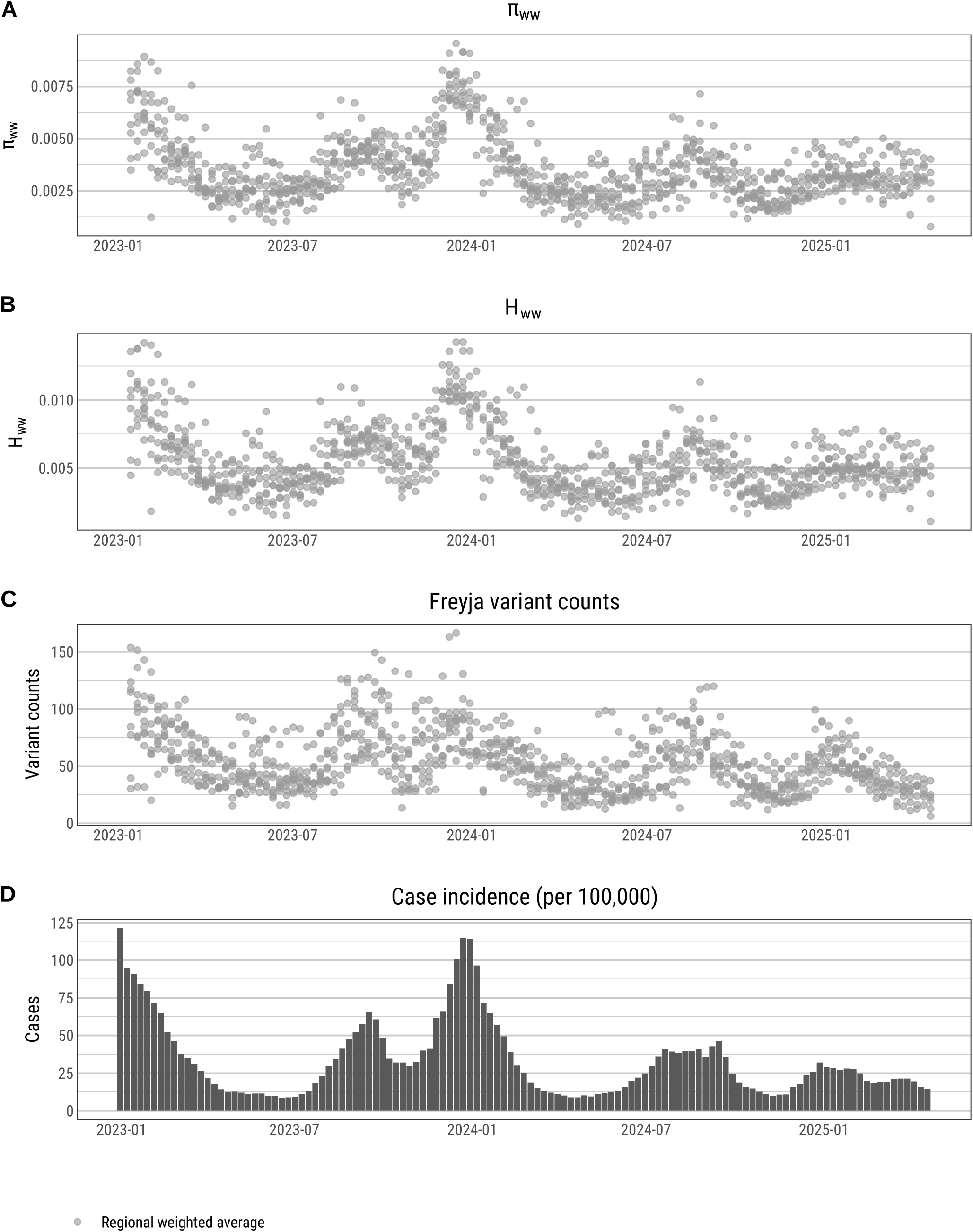
A) Regional population weighted average *π*_s_ estimates for S1 NTD region per county, per week. B) Regional population weighted average H_s_ estimates for the S1 NTD region per county, per week. C) Regional population weighted average number of Freyja variant counts per county per week. D) Weekly statewide total cases per 100,000 population.

**Figure S4:**
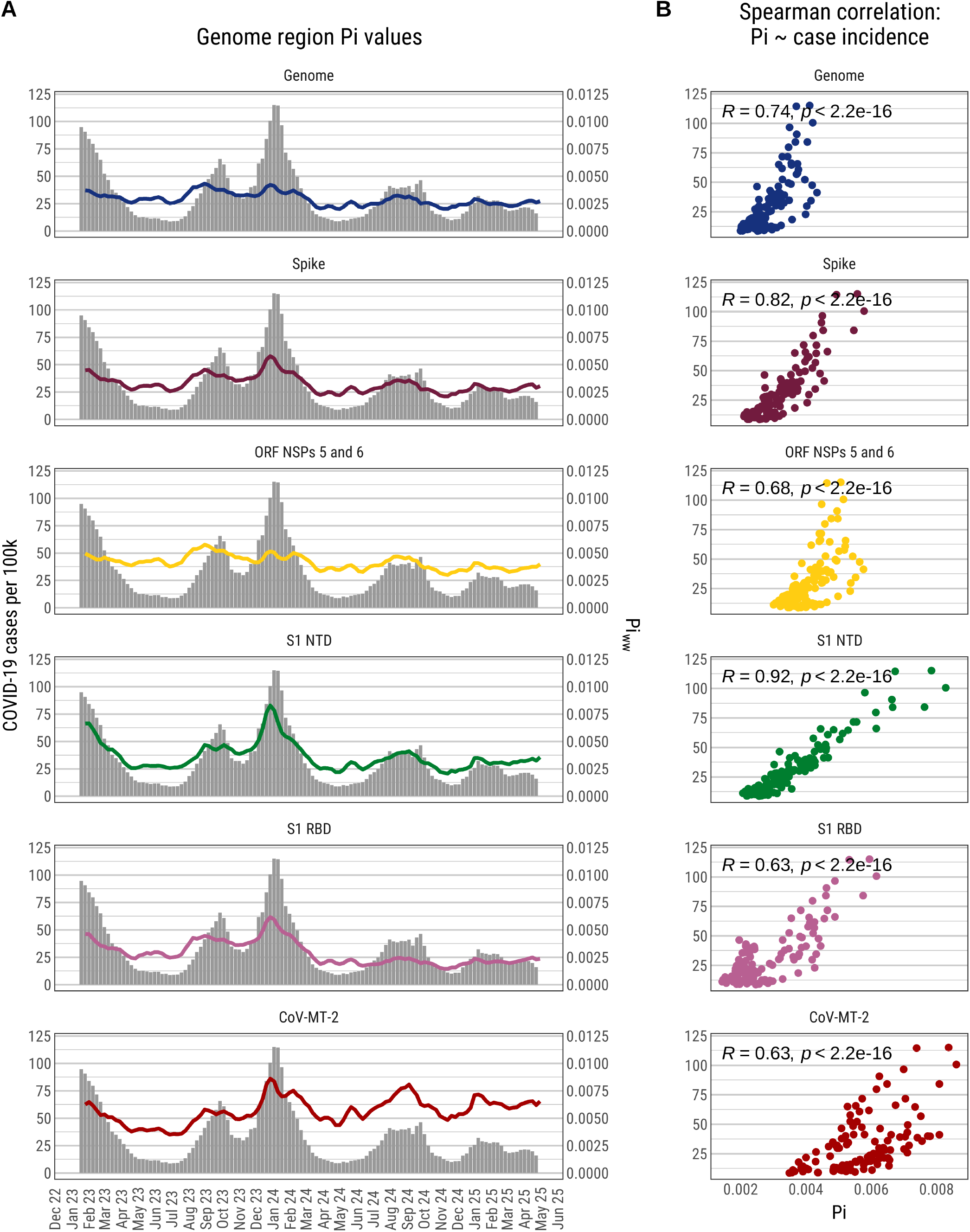
Time series plots for each genome region of interest *π*_s_ values and COVID-19 case incidence. Each region correlated with incidence, including genome-wide diversity.

**Figure S5:**
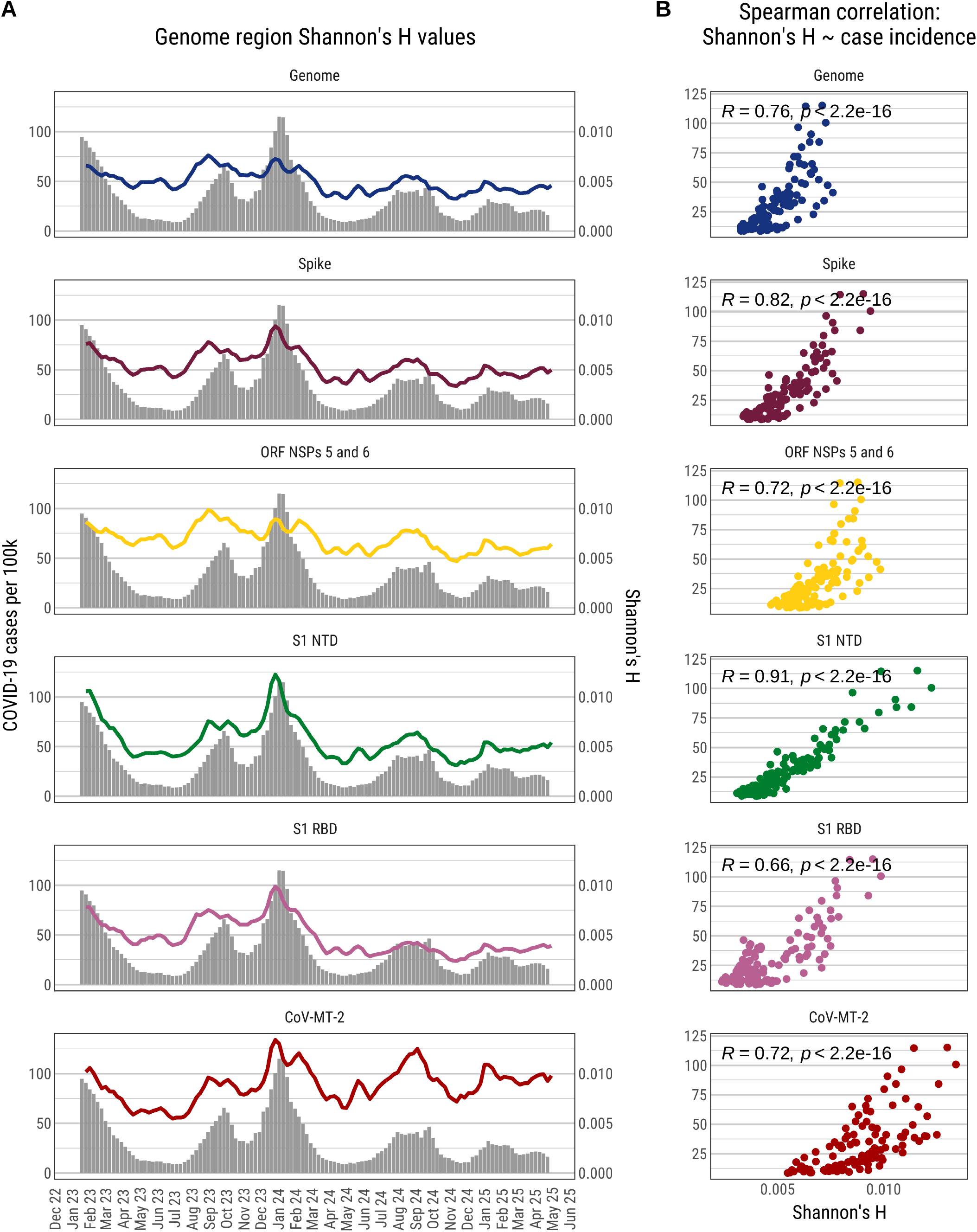
Time series plots for each genome region of interest H_s_ values and COVID-19 case incidence. Each region correlated with incidence, including genome-wide diversity.

**Figure S6:**
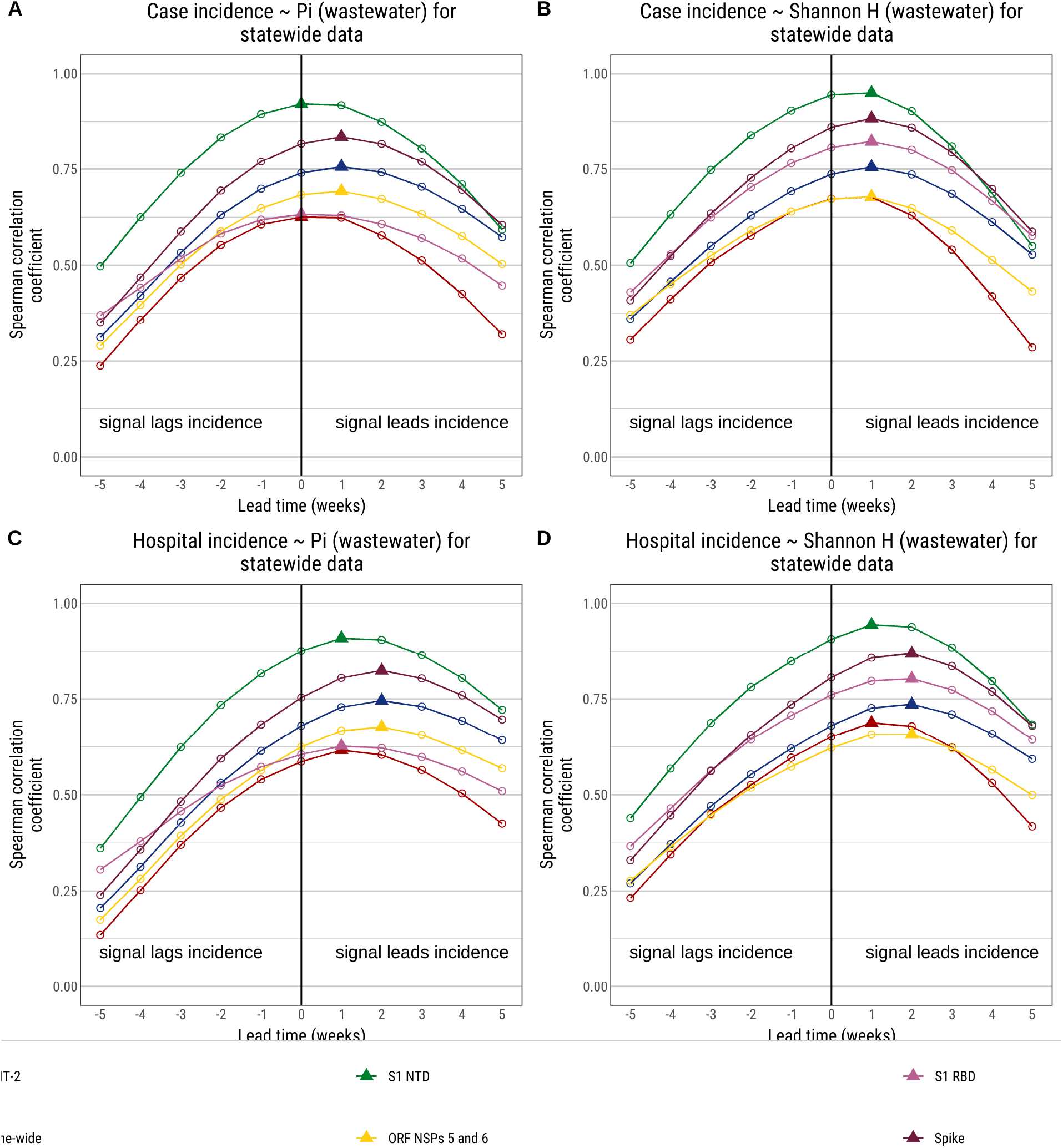
A) Lag and lead Spearman correlations for each genome region for *π*_s_ and case incidence. B) Lag and lead Spearman correlations for each genome region for H_s_ and case incidence. C) Lag and lead Spearman correlations for each genome region for *π*_s_ an hospitalization incidence. D) Lag and lead Spearman correlations for each genome region for H_s_ and hospitalization incidence.

**Table S3:** Statewide generalized least squares model results for S1 NTD. Model has a correction for time series data (AR1), coefficients are standardized, and the outcome, COVID-19 case incidence, is on its original scale. Concentration is log transformed.

| Model | Variable/Metric | Estimate | Standard Error (SE) | t value | P value |
| --- | --- | --- | --- | --- | --- |
| Full model | Intercept | 32.131 | 0.624 | 51.46*** | <0.0001 |
|  | S1 NTD Pi | 20.206 | 0.999 | 20.23*** | <0.0001 |
|  | SARS-CoV-2 concentration | 3.790 | 0.840 | 4.51*** | <0.0001 |
|  | Mean sample depth of read | -0.696 | 1.338 | -0.52 | 0.61 |
|  | Number of samples sequenced | 1.529 | 0.870 | 1.76 | 0.09 |
|  | Number of samples collected | -1.060 | 0.819 | -1.29 | 0.20 |
|  | Mean genome coverage | -0.134 | 0.807 | -0.17 | 0.87 |
|  | n | 119.000 |  |  |  |
|  | Efron R2 | 0.921 |  |  |  |
|  | AIC | 796.302 |  |  |  |
| S1 NTD model | Intercept | 32.131 | 0.684 | 46.98*** | 0.00 |
|  | S1 NTD Pi | 22.406 | 0.687 | 32.63*** | 0.00 |
|  | n | 119.000 |  |  |  |
|  | Efron R2 | 0.901 |  |  |  |
|  | AIC | 819.821 |  |  |  |
| Concentration model | Intercept | 32.131 | 1.716 | 18.72*** | 0.00 |
|  | SARS-CoV-2 concentration | 14.478 | 1.724 | 8.4*** | 0.00 |
|  | n | 119.000 |  |  |  |
|  | Efron R2 | 0.376 |  |  |  |
|  | AIC | 1,035.160 |  |  |  |

**Table S4:** County generalized mixed model results for S1 NTD. Model has a negative binomial distribution. Model has a correction for time series data (AR1), coefficients are standardized, and the outcome, COVID-19 case incidence, is on its original scale. Concentration is log transformed. County is a random effect in the model.

| Variable/Metric | Estimate | Standard Error (SE) | t value | p_value |
| --- | --- | --- | --- | --- |
| Intercept | 3.63700 | 0.05090 | 71.51*** | <0.0001 |
| S1 NTD Pi | 0.09680 | 0.01150 | 8.39*** | <0.0001 |
| SARS-CoV-2 concentration | 0.08510 | 0.01270 | 6.69*** | <0.0001 |
| Mean sample depth of read | -0.00176 | 0.00885 | -0.2 | 0.84 |
| Number of samples sequenced | 0.00977 | 0.00824 | 1.19 | 0.24 |
| Number of samples collected | 0.00974 | 0.02120 | 0.46 | 0.65 |
| Mean genome coverage | -0.00282 | 0.00592 | -0.48 | 0.63 |
| n | 4,397.00000 |  |  |  |
| Efron R2 | 0.99600 |  |  |  |
| AIC | 36,881.97000 |  |  |  |

**Table S5:** Regional generalized mixed model results for S1 NTD. Model has a negative binomial distribution. Model has a correction for time series data (AR1), coefficients are standardized, and the outcome, COVID-19 case incidence, is on its original scale. Concentration is log transformed. Region is a random effect in the model.

| Variable/Metric | Estimate | Standard Error (SE) | t value | p_value |
| --- | --- | --- | --- | --- |
| Intercept | 3.6740 | 0.1540 | 23.89*** | <0.0001 |
| S1 NTD $\pi_i$ | 0.1290 | 0.0210 | 6.16*** | <0.0001 |
| SARS-CoV-2 concentration | 0.1040 | 0.0251 | 4.14*** | <0.0001 |
| Mean sample depth of read | 0.0120 | 0.0140 | 0.85 | 0.39 |
| Number of samples sequenced | 0.0172 | 0.0113 | 1.51 | 0.13 |
| Number of samples collected | 0.0183 | 0.0292 | 0.63 | 0.53 |
| Mean genome coverage | -0.0088 | 0.0100 | -0.88 | 0.38 |
| n | 1,007.0000 |  |  |  |
| Efron R2 | 0.9960 |  |  |  |
| AIC | 7,388.3160 |  |  |  |

**Table S6:** Granger causality results. Each diversity measure (x) is tested to see if a one week lagged value would predict case incidence and hospitalization incidence (y). Granger causality also tests to determine if y would predict x. Our findings show that lagged diversity measures (x) are always strong predictors of y but lagged y values do not predict x. Thus, the diversity measures are predictive of clinical data.

| Diversity measure (x) | Outcome variable (y) | Model | F statistic | P value |
| --- | --- | --- | --- | --- |
| S1 NTD $\pi_{ww}$ | Case incidence | x predicts y | 52.3600 | <0.001*** |
| S1 NTD $\pi_{ww}$ | Case incidence | y predicts x | 0.1230 | 0.726 |
| S1 NTD $H_{ww}$ | Case incidence | x predicts y | 44.9840 | <0.001*** |
| S1 NTD $H_{ww}$ | Case incidence | y predicts x | 0.3410 | 0.561 |
| Freyja variant count | Case incidence | x predicts y | 13.9640 | <0.001*** |
| Freyja variant count | Case incidence | y predicts x | 3.0230 | 0.0848 |
| S1 NTD $\pi_{ww}$ | Hospitalization incidence | x predicts y | 52.1770 | <0.001*** |
| S1 NTD $\pi_{ww}$ | Hospitalization incidence | y predicts x | 0.0117 | 0.914 |
| S1 NTD $H_{ww}$ | Hospitalization incidence | x predicts y | 47.7970 | <0.001*** |
| S1 NTD $H_{ww}$ | Hospitalization incidence | y predicts x | 0.0709 | 0.791 |
| Freyja variant count | Hospitalization incidence | x predicts y | 20.0520 | <0.001*** |
| Freyja variant count | Hospitalization incidence | y predicts x | 0.6020 | 0.439 |

**Table S7:** Model for predicting *π*ww from sample depth. Variance explained for *π*s by sample depth is 14 percent.

| variable | Est | Std. Error | t value2 | Pr(> \|t\|) |
| --- | --- | --- | --- | --- |
| Intercept | 0.002930 | 0.0000130 | 226.21*** | <0.0001 |
| Sample mean depth | 0.000534 | 0.0000129 | 41.26*** | <0.0001 |
| n | 10,375.000000 |  |  |  |
| Efron R2 | 0.141000 |  |  |  |

**Figure S7:**
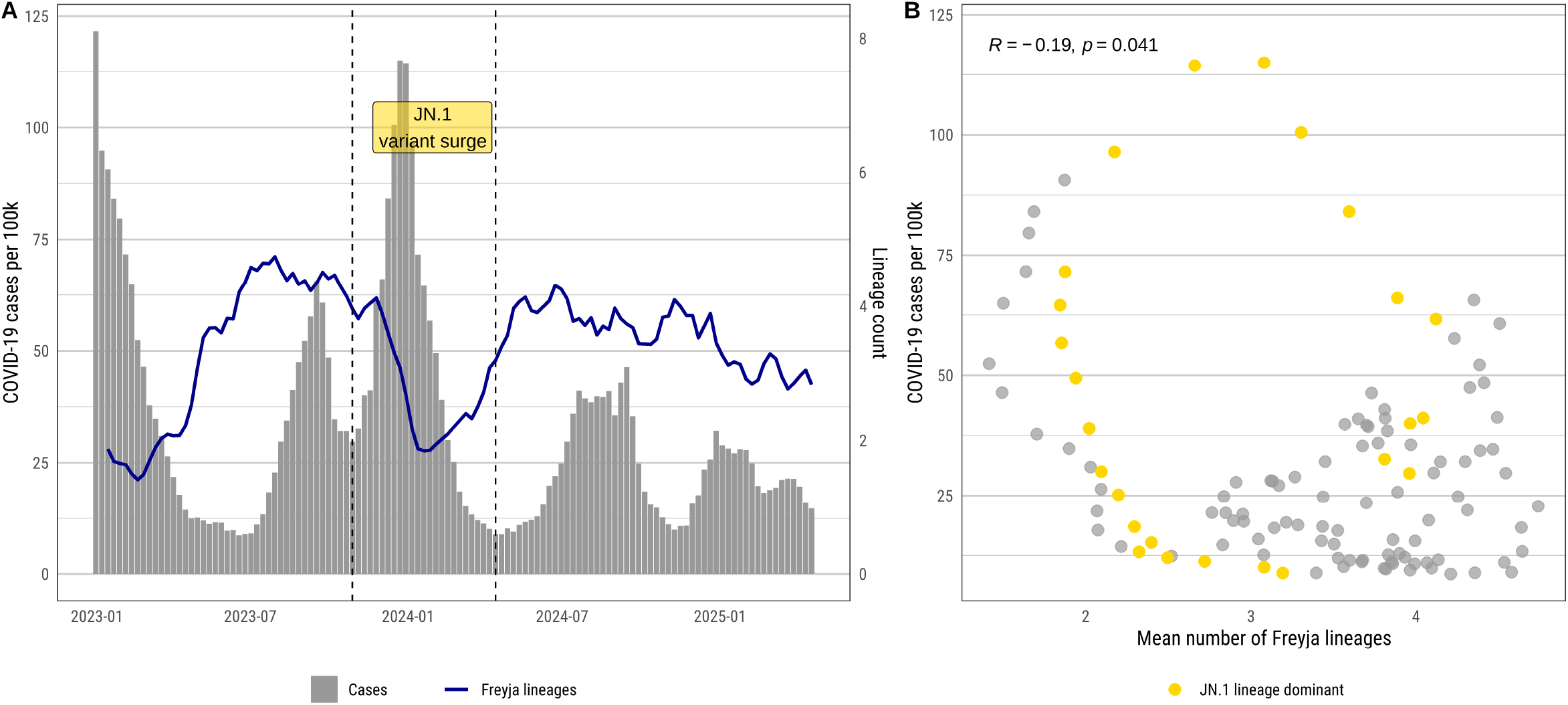
Number of virus lineages when an abundance threshold of five percent is applied. A) Freyja lineage counts with a five percent threshold applied and COVID-19 case incidence. B) Scatterplot and Spearman correlation coefficient for Freyja lineage count and COVID-19 case incidence.

**Figure S8:**
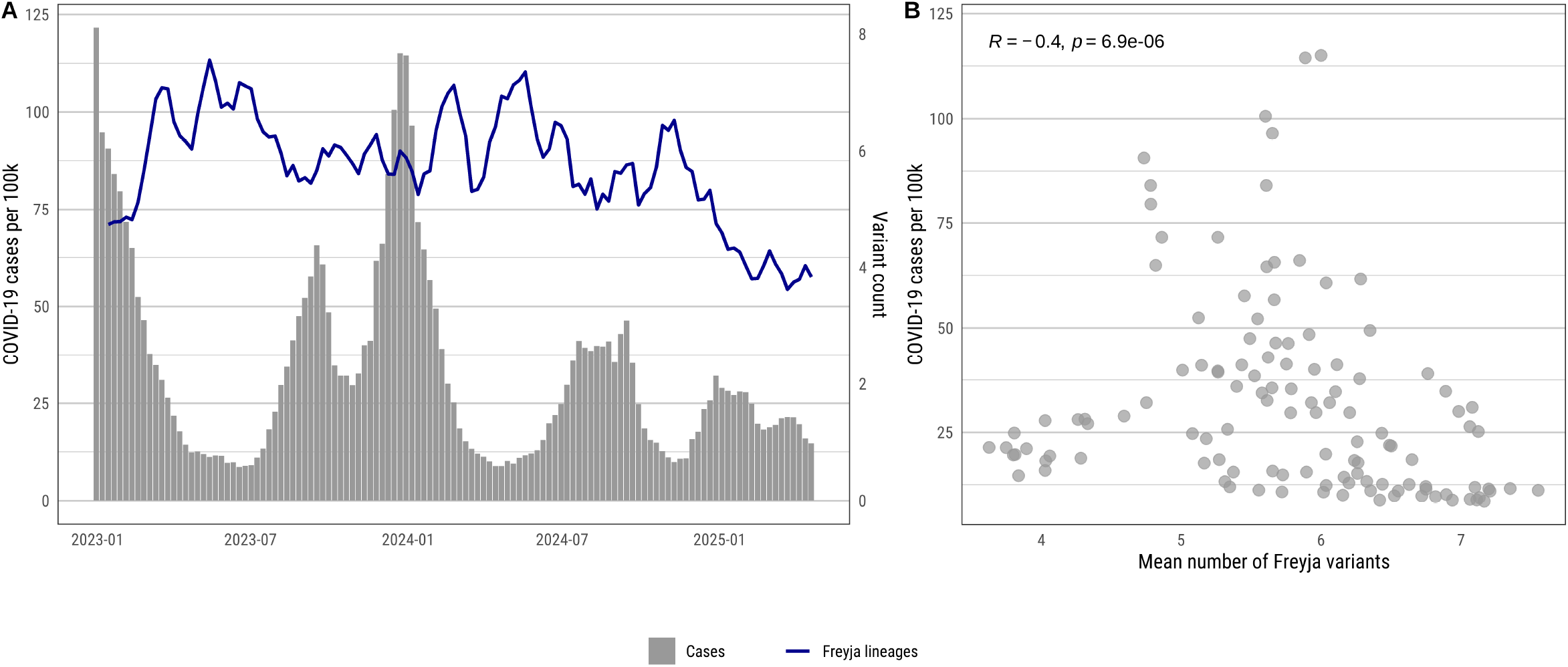
A) Freyja variant counts with a 5% threshold applied and COVID-19 case incidence. B) Scatterplot and Spearman correlation of Freyja variant counts and COVID-19 case incidence.

**Figure S9:**
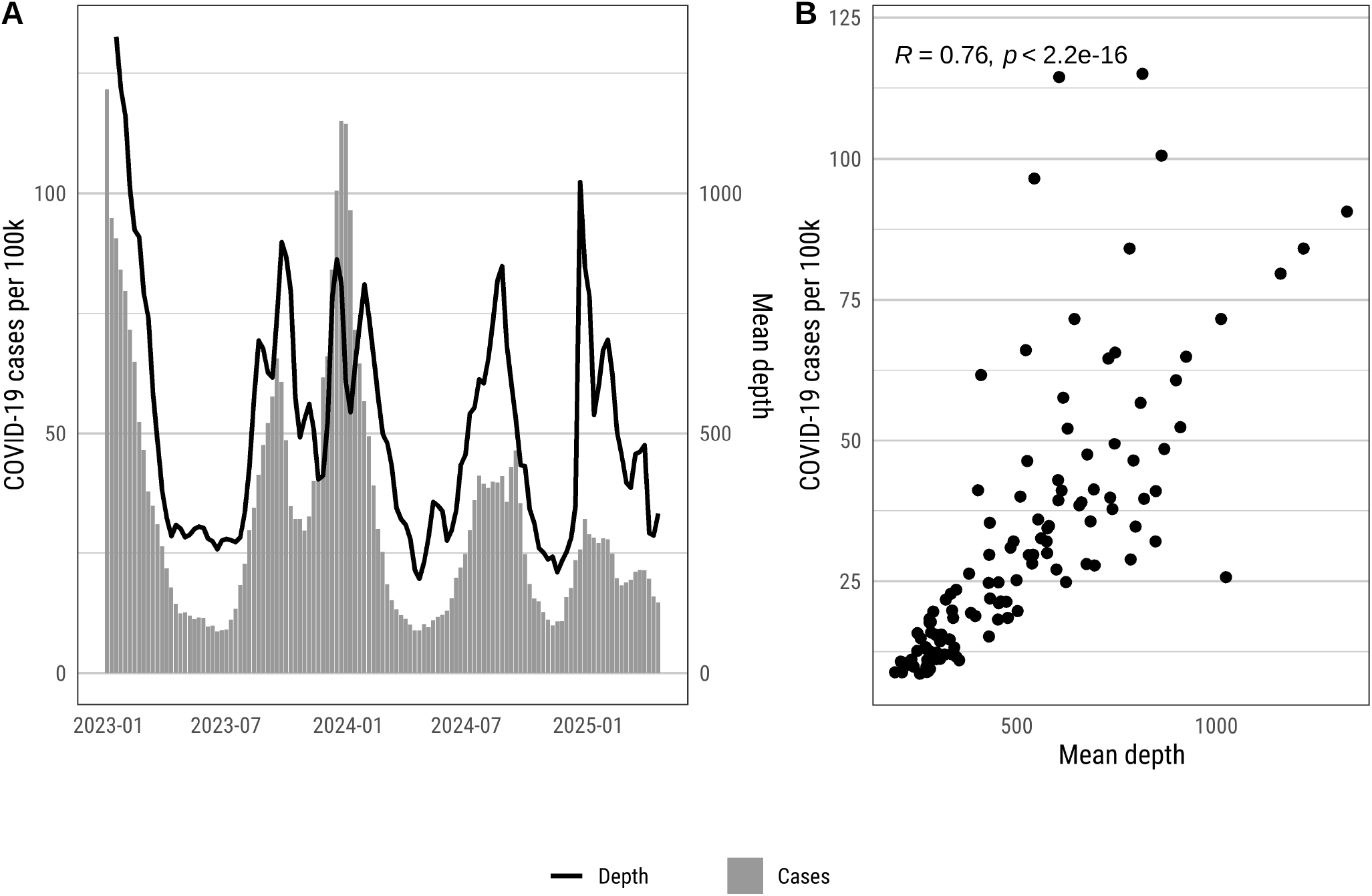
A) Population weighted mean depth of read and COVID-19 cases over time. B) Spearman correlation for mean depth and COVID-19 cases. As COVID-19 cases increase, overall depth per sample increases.

**Figure S10:**
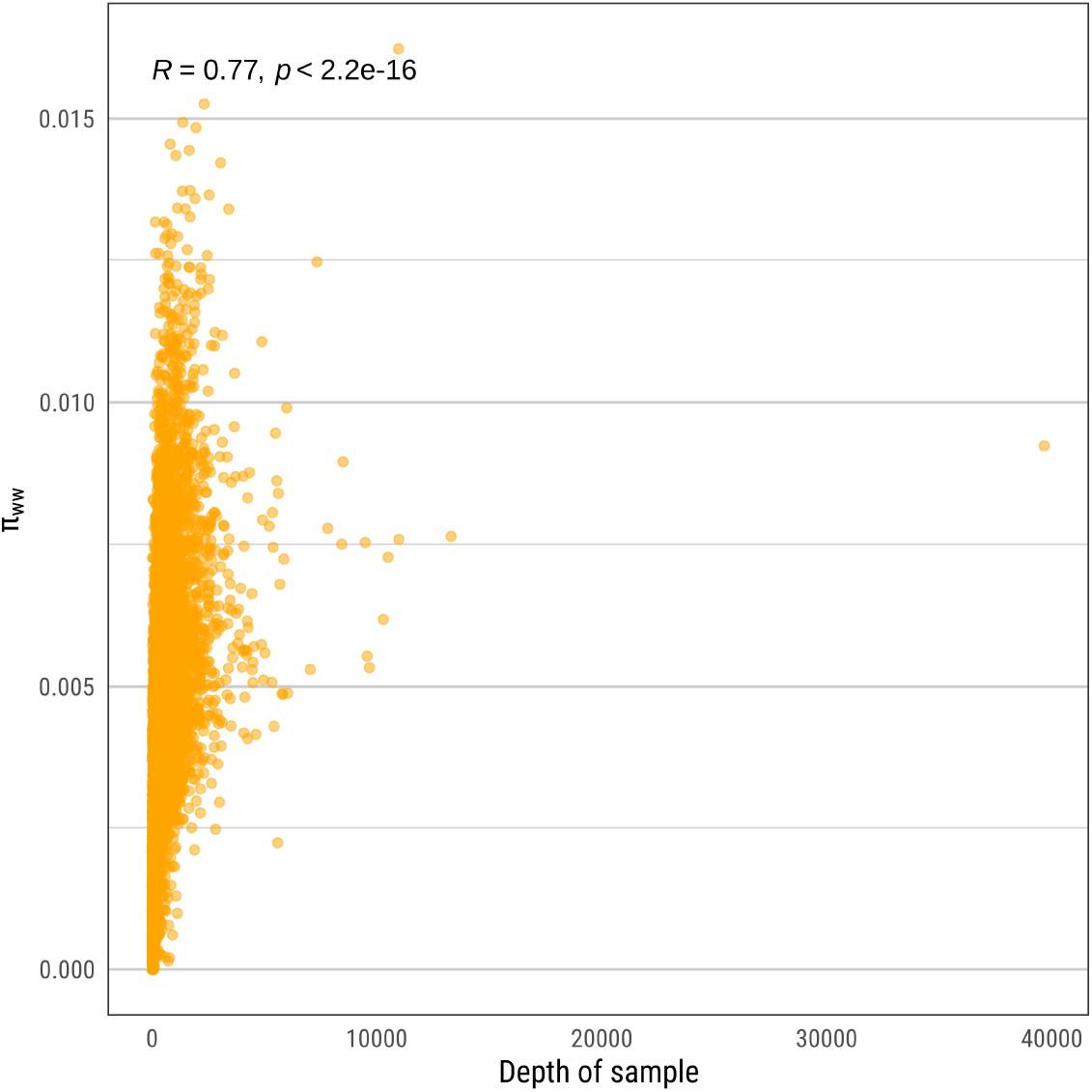
Scatterplot and Spearman correlation for depth per sample and *π*_s_.

**Figure S11:**
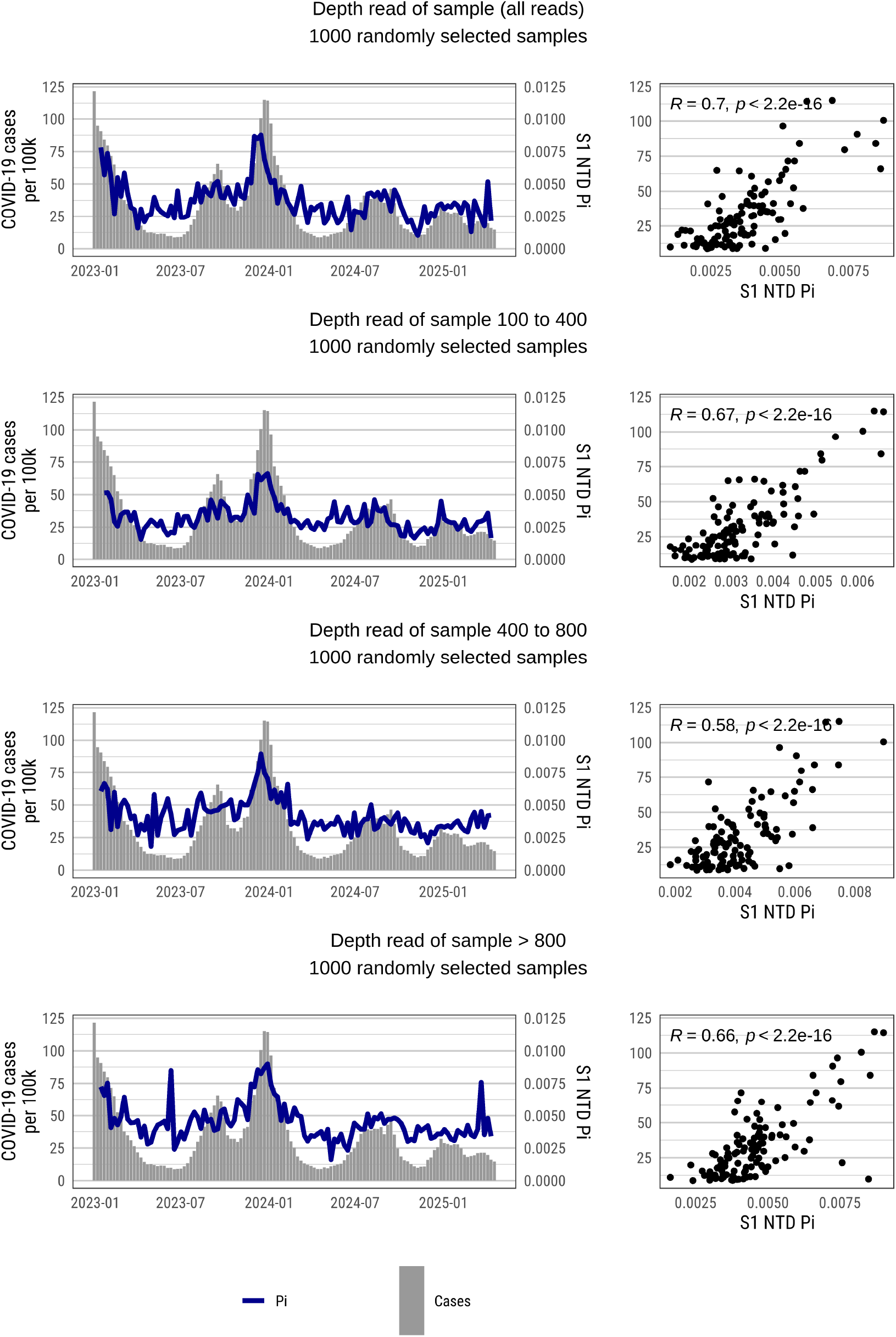
Random samples of wastewater sequences of equal read depth. Randomly sampling the dataset for equal read depth did not change the overall findings with the mean S1 NTD *π*_s_ having the same relationship with cases regardless of read depth.

**Table S8:**
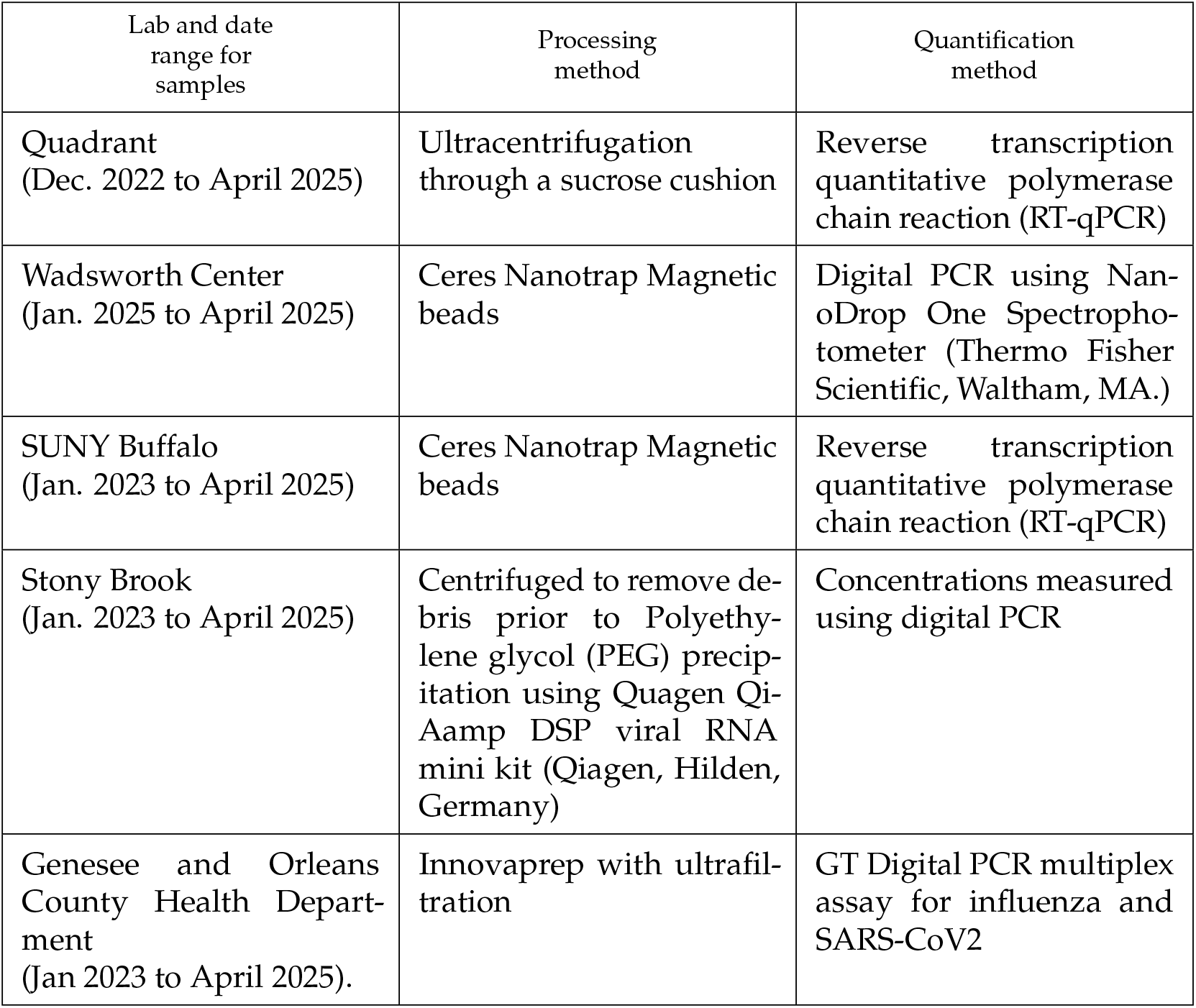
Table S8: Methods used by regional PCR labs.

| Lab and date range for samples | Processing method | Quantification method |
| --- | --- | --- |
| Quadrant (Dec. 2022 to April 2025) | Ultracentrifugation through a sucrose cushion | Reverse transcription quantitative polymerase chain reaction (RT-qPCR) |
| Wadsworth Center (Jan. 2025 to April 2025) | Ceres Nanotrap Magnetic beads | Digital PCR using NanoDrop One Spectrophotometer (Thermo Fisher Scientific, Waltham, MA.) |
| SUNY Buffalo (Jan. 2023 to April 2025) | Ceres Nanotrap Magnetic beads | Reverse transcription quantitative polymerase chain reaction (RT-qPCR) |
| Stony Brook (Jan. 2023 to April 2025) | Centrifuged to remove debris prior to Polyethylene glycol (PEG) precipitation using Quagen QiAamp DSP viral RNA mini kit (Qiagen, Hilden, Germany) | Concentrations measured using digital PCR |
| Genesee and Orleans County Health Department (Jan 2023 to April 2025). | Innovaprep with ultrafiltration | GT Digital PCR multiplex assay for influenza and SARS-CoV2 |

**Table S9:** Descriptive statistics for concentration data.

| PCR Lab | Number of sampling sites | Number of samples | First collection week | Last collection week | Mean | Min | Median | Max | SD |
| --- | --- | --- | --- | --- | --- | --- | --- | --- | --- |
| GO Health | 7 | 196 | 2023-01-01 | 2025-04-20 | 6.8 | 1.5 | 3.5 | 130 | 11 |
| Quadrant | 157 | 9,276 | 2023-01-01 | 2025-04-20 | 28.0 | 0.0 | 3.5 | 99,000 | 870 |
| SUNY Buffalo | 22 | 1,335 | 2023-01-01 | 2025-04-20 | 1,400.0 | 0.0 | 520.0 | 89,000 | 3,700 |
| SUNY Stony Brook | 18 | 1,323 | 2023-01-01 | 2025-04-20 | 26.0 | 0.0 | 13.0 | 1,100 | 48 |
| Wadsworth Center | 61 | 236 | 2024-12-22 | 2025-04-20 | 76.0 | 0.0 | 41.0 | 1,300 | 130 |

**Table S10:**
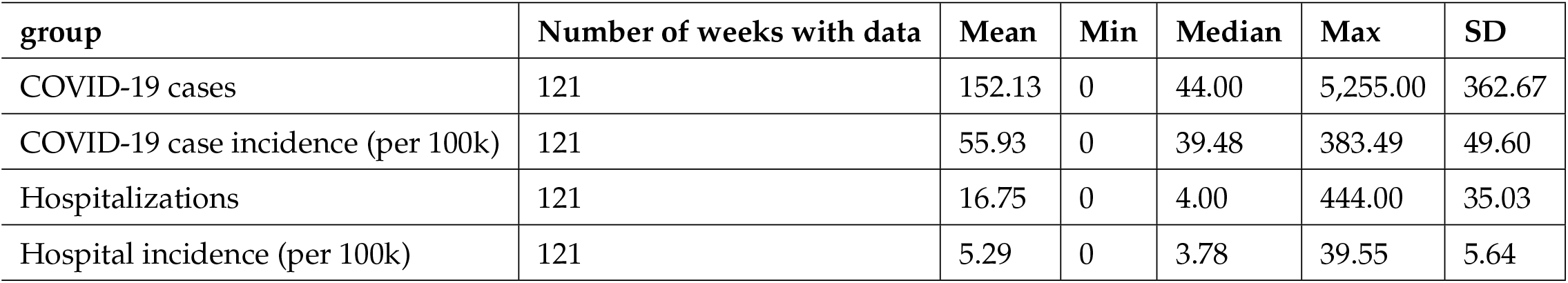
Descriptive statistics for COVID-19 clinical data.

| group | Number of weeks with data | Mean | Min | Median | Max | SD |
| --- | --- | --- | --- | --- | --- | --- |
| COVID-19 cases | 121 | 152.13 | 0 | 44.00 | 5,255.00 | 362.67 |
| COVID-19 case incidence (per 100k) | 121 | 55.93 | 0 | 39.48 | 383.49 | 49.60 |
| Hospitalizations | 121 | 16.75 | 0 | 4.00 | 444.00 | 35.03 |
| Hospital incidence (per 100k) | 121 | 5.29 | 0 | 3.78 | 39.55 | 5.64 |

**Table S11:** Genome regions and base pair positions that had diversity values calculated.

| **Genome region/product** | **Product or product feature** | **Base pair start** | **Base pair end** |
| --- | --- | --- | --- |
| ORF 1a region | NSP 5 | 10063 | 10954 |
|  | NSP 6 | 10972 | 11842 |
| ORF 1b region | COV-Methyltr-2 | 20661 | 21549 |
| Spike Protein | S1 N-terminal domain (NTD) | 21598 | 22474 |
|  | S1 receptor binding domain (RBD) | 22516 | 23185 |

**Figure S12:**
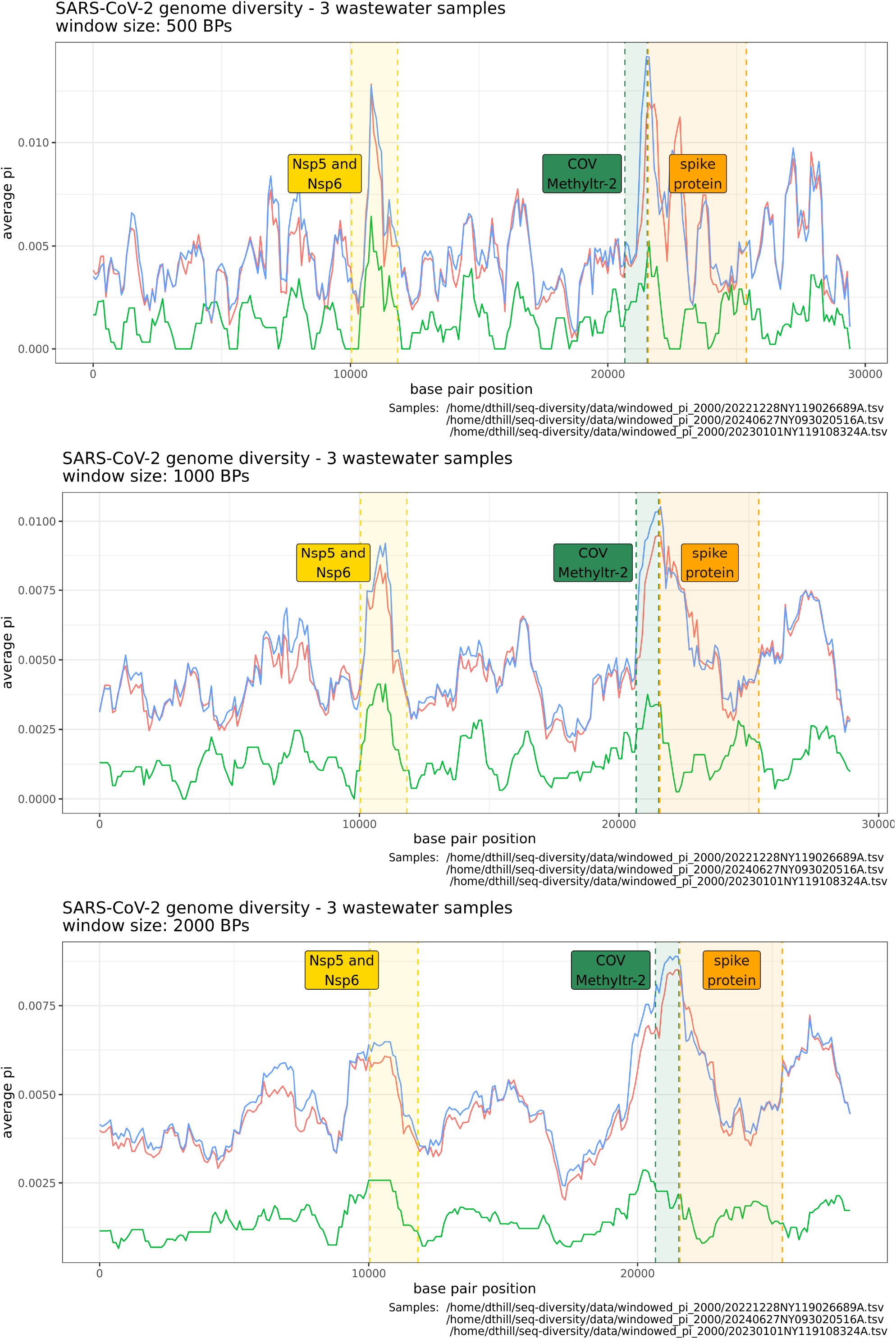
Windowed diversity values for three example samples calculated with three different windows for comparison (500, 1000, and 200 bps). Each window size resulted in similar findings for high diversity in the regions of interest.

**Figure S13:**
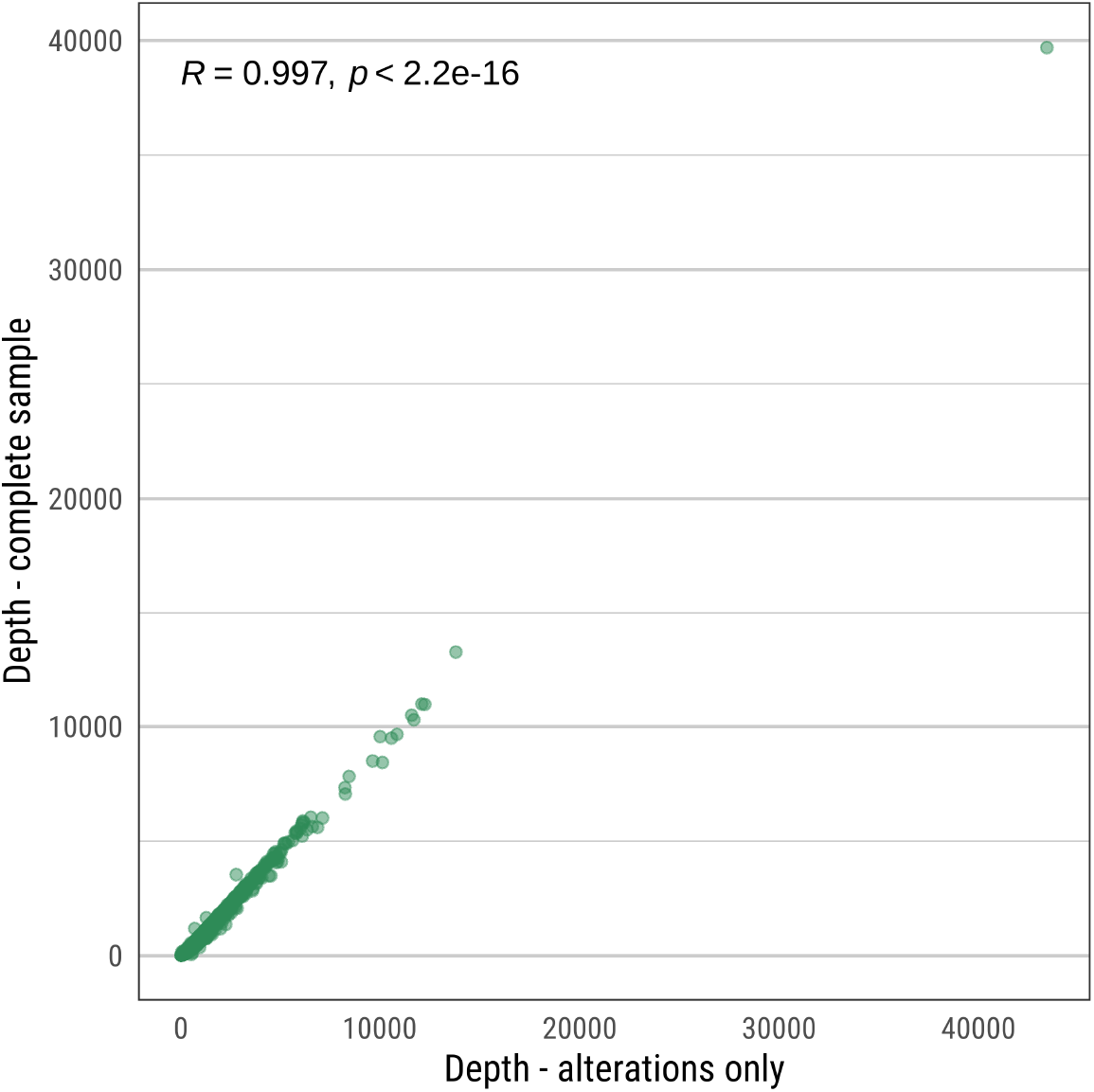
Scatterplot and Spearman correlation for depth of read per sample and depth for variants (base pair changes) only from the Freyja output.

